# Compound climate extremes, socioeconomic conditions and human mobility influence dengue dynamics heterogeneously across Vietnam

**DOI:** 10.64898/2026.09.19.26363468

**Authors:** Chloe Fletcher, Sophie Belman, Kien Quoc Do, Quang Duy Pham, Thi Thanh Thao Nguyen, Rory Gibb, Phan Trong Lan, Tran Cong Tu, Nguyen Hai Tuan, Daniela Lührsen, Gina Tsarouchi, Quillon Harpham, Felipe J. Colón-González, John Palmer, Vu Sinh Nam, Rachel Lowe

## Abstract

Dengue presents a major public health challenge in Vietnam, driven by biological, behavioural, and environmental factors. Compound climate extremes, such as sequential hydrometeorological events, can influence dengue risk yet remain understudied. We evaluated the effects of compound climate extremes, socioeconomic conditions, and human mobility on dengue relative risk across 670 districts in Vietnam over 20 years, stratifying by eight subregions spanning emerging to endemic transmission. Dengue risk was greatest following dry-then-wet conditions in seven subregions. Higher temperatures increased risk across North and Central Vietnam, but had limited effect in the South. Mobility associations followed an urban-rural gradient, with increased risk in highly rural districts where residents travelled more frequently to fewer destinations, and in urban districts with more visitors and dispersed outgoing mobility. Stratifying climatic and socioeconomic effects by subregion improved predictive skill at lead times of 1 to 6 months over unstratified and baseline models, with substantial spatial variation. Accounting for compound extremes across distinct spatial contexts could strengthen disease early warning systems in Vietnam and beyond.

## Introduction

Dengue fever is a major public health challenge globally, with Southeast Asia among the most affected regions (World Health Organization, 2024). Historically, Vietnam has borne a high share of this burden, reporting 2.5 million infections between 1980 and 2010 (Wartel et al., 2016). A recent meta-analysis estimated national dengue seroprevalence to be 41.0% (95% CI: 30.5-51.4%) with regional heterogeneity ranging from 3.1% to 77.1% (Le et al., 2026). These spatial disparities reflect distinct epidemiological profiles across Vietnam’s latitudinal gradient. In the subtropical North, dengue is an emerging disease characterised by intermittent outbreaks, whereas in the tropical Central and South regions it is largely endemic or hyperendemic (Colón-González et al., 2021). As the drivers of dengue emergence often differ from those sustaining transmission (Gibb et al., 2023), modelling frameworks require sufficient flexibility and complexity to capture these varied dynamics.

Climatic conditions influence dengue transmission across Vietnam by shaping both vector ecology and human behaviour (Vu et al., 2014). *Aedes aegypti*, the primary global vector, thrives in warmer, urban and domestic environments typical of Central and Southern Vietnam (Duong et al., 2022). However, the more cool-tolerant *Aedes albopictus*, which predominates in Northern Vietnam, is native to Southeast Asia with demonstrated vector competence across the continent (Doeurk et al., 2024; Joanne et al., 2017; Kobayashi et al., 2023). As these species exhibit specific thermal optima (Kramer et al., 2026; Mordecai et al., 2019), the meteorological drivers of dengue likely vary across this ecological gradient. Additionally, human adaptation to weather, such as domestic water storage during dry periods, increases transmission risk (Schmidt et al., 2011; D. Wang et al., 2026). Previous research evaluating the nonlinear and delayed effects of climate on dengue in Vietnam found increased risks with drought (lagged 5 months), heavy rainfall (lagged 1 month), warmer mean temperature of the coolest month (annually), and monthly mean temperatures ranging 23-33°C (lagged 1 month) (Gibb et al., 2023). These climatic drivers are consistent with other studies in endemic settings (Lowe et al., 2018, 2021). Few frameworks have evaluated how these climatic conditions compound to elevate dengue risk, and whether that response differs systematically across emerging and endemic regimes.

Compound events are combinations of multiple climatic hazards, such as extremes, that contribute to societal or environmental risk (Zscheischler et al., 2018). These events can be grouped into four types: (1) preconditioned, where a pre-existing condition or hazard amplifies the impact of a subsequent hazard, (2) multivariate, where multiple hazards coincide locally to form a combined impact, (3) temporally compounding, where locally successive hazards form a combined impact, and (4) spatially compounding, where hazards coincide across connected locations to form a combined impact (Zscheischler et al., 2020). Most compound event analyses of dengue have examined multivariate events. Concurrent warm and wet conditions increased dengue relative risk compared to moderate or isolated events in 48 locations across Malaysia, Singapore, Sri Lanka, and Thailand (Y. Wang et al., 2024). Across seven Southeast Asian countries at the first administrative level, including Vietnam, compound drought-heatwave events carried a lower maximum relative risk of dengue than droughts or heatwaves alone (D. Wang et al., 2026). Furthermore, causal analysis identified drought and drought-heatwave events as key drivers in urbanised areas, but not in rural areas. Sequential events have received less attention. In Barbados, the greatest dengue outbreak risk emerged after successive drought (lagged 5 months), heat (lagged 3 months), and extreme rainfall (lagged 1 month) (Fletcher et al., 2025). The boundary between compound event types is often unclear in practice without a definitive mechanistic pathway (Zscheischler et al., 2020). At present, the effects of compound climate extremes on dengue risk have not been quantified at a fine spatial scale in Vietnam.

Dengue transmission is further impacted by other anthropogenic factors. Urbanisation, land-use change, and disparities in water and sanitation infrastructure influence human behaviour and vector proliferation (Gibb et al., 2023; Lee et al., 2021; Lowe et al., 2021). Spatial connectivity and mobility are also major determinants. By introducing dengue virus to immunologically susceptible populations, human movement can catalyse localised epidemics, particularly in emerging settings (Harish et al., 2024; Salje et al., 2021). Many modelling frameworks account for connectivity using adjacency matrices, assuming that disease spreads stepwise to neighbouring areas (Colón-González et al., 2021; Gibb et al., 2023; Lowe et al., 2021). However, mobility is also driven by economic hubs, transit networks, available amenities, and tourism, which can connect geographically distant populations (Belman et al., 2024; Kiang et al., 2021; Salje et al., 2021). Alternative representations of spatial connectivity may therefore capture more realistic pathways of dengue spread.

Translating this complex interplay of climatic, socioeconomic and mobility drivers into predictive models can support public health preparedness and response. Early warning systems mitigate epidemic risk by providing decision-makers with actionable lead time to allocate clinical resources, conduct community outreach, and deploy targeted vector control before an outbreak occurs (Alcayna et al., 2025; Díaz et al., 2024). In Vietnam, a forecasting framework utilising lagged meteorological covariates demonstrated skill in predicting province-level dengue incidence 1 to 6 months ahead (Colón-González et al., 2021). A subsequent framework modelled dengue dynamics at the district level, but did not systematically evaluate or optimise predictive skill at operational lead times (Gibb et al., 2023). Building on these frameworks, fine-scale dengue predictability could be enhanced by accounting for the interactive effects of compound climate extremes, and shifting from national-scale to spatially-stratified parameter estimates to capture distinct dynamics across diverse settings.

This study presents a Bayesian spatiotemporal mixed-effects modelling framework to evaluate dengue associations and predictors across 670 districts in Vietnam over a 20-year period. We quantify the effects of compound climate extremes on dengue relative risk using interactions between long-lag and short-lag drought indicators. We further assess other climatic, oceanic, socioeconomic, and mobility covariates as additive terms, as well as adjacency and movement matrices to characterise spatial dependence. We utilise random slopes to stratify interactive and additive effects across different spatial classifications, including geographic regions, climatic zones, urbanisation levels, and elevation bands, to account for spatially distinct dengue dynamics. Finally, we evaluate model predictive skill at operational lead times of 1 to 6 months in advance across various spatial scales. This approach aims to enhance our understanding of dengue to support early warning initiatives at a fine spatial scale in Vietnam. This framework is adaptable in other settings for modelling spatially varying climate-sensitive disease risk.

## Results

### Dengue incidence and climatic conditions vary along Vietnam’s latitudinal gradient

Surveillance records reported 1,991,432 clinically diagnosed (i.e. confirmed or suspected) cases of dengue across 670 districts between May 2001 and April 2021, representing a mean annual incidence of 113 cases per 100,000 population. Dengue incidence was distributed heterogeneously across the country (Figure 1A), including subregions in the North (Northwest, Northeast, Red River Delta), Central (North Central, South Central Coast, Central Highlands) and South (Southeast, Mekong River Delta) (Figure 1B). The highest mean annual incidence was found in the Southeast with 223 cases per 100,000 population, followed by the South Central Coast with 192, Mekong River Delta with 186, and Central Highlands with 137. The lowest mean annual incidence was found in the Northwest and Northeast with 3 and 5 cases per 100,000 population, respectively. This transmission gradient aligns with elevation (Figure 1C), climate classification (Figure 1D), and levels of urbanisation (Figure S1). Dengue cases exhibited a consistent seasonality throughout the country, although regional case magnitudes varied each year (Figure S2A).

**Figure 1.**
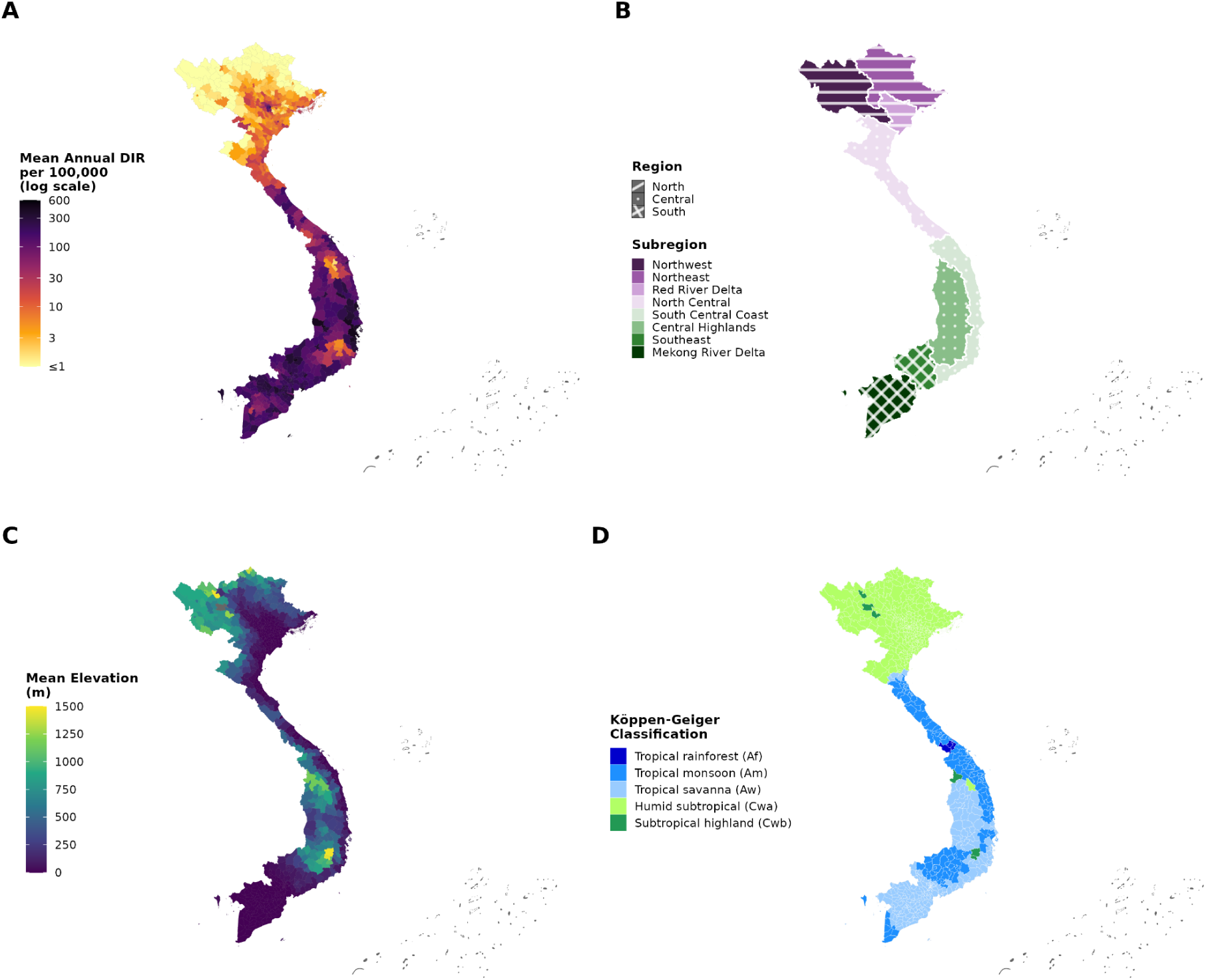
Spatial distribution of dengue incidence, elevation, climate classification, and administrative regions across Vietnam. (A) Mean annual dengue incidence rate (DIR) per 100,000 inhabitants per district between May 2001 and April 2021. (B) The three regions (shading) and eight subregions (colour) of Vietnam. (C) Mean district elevation (m) derived from the Copernicus GLO-90 Digital Elevation Model. (D) Modal Köppen-Geiger climate classification per district for the 1991-2020 reference period (Beck et al., 2023). Districts represent merged Global Administrative Areas (GADM) boundaries (n = 670), with some island district values greyed out due to missing data points.

There are distinct climatic patterns across the country (Figure S2C-F). Mean temperature over rolling 6-month windows averaged 22.9°C (17.4-27.5°C) in the North, 24.4°C (20.9-27.6°C) in the Central region, and 26.9°C (25.6-28.3°C) in South Vietnam (Figure S2C), yet the timing of temperature anomalies was largely consistent across regions (Figure S2D). Monthly precipitation totals, averaged over the same rolling 6-month windows, were similar in magnitude across regions – 156.2 mm/month (33.3-299.0 mm/month) in the North, 164.1 mm/month (60.4-304.3 mm/month) in the Central region, and 166.6 mm/month (20.4-317.1 mm/month) in the South – but differed in seasonal timing (Figure S2E). The 6-month Standardised Precipitation Evapotranspiration Index (SPEI-6) was heterogeneous in space and time, with dry and wet extremes concurrent across different regions (Figure S2F).

We first identified the best-fitting model specification through goodness-of-fit metrics and the assessment of credible effect sizes (i.e. 95% credible interval excludes zero). We report the estimated effects of the climatic, socioeconomic, and mobility covariates on dengue relative risk across Vietnam. We then quantified predictive skill under rolling-origin cross-validation with an expanding window from May 2016 to April 2021 at 1-to 6-month lead times. We present predictive metrics aggregated across all district-months, and subsequently at subregional and district-level scales to evaluate skill for potential early warning initiatives.

### Compound climatic and socioeconomic associations with dengue are spatially heterogeneous

We designed and followed a stepwise model development framework (Figure S3), fitting Bayesian hierarchical mixed-effects models on 20 years of district-level data to identify associations between covariates and dengue incidence. The best-fitting model accounted for compound climatic extremes via a two-way interaction between the SPEI-6 at both a long lag (5 months) and short lag (0 months), along with additive terms for the absolute 6-month mean temperature (lagged 0 months), the Dipole Mode Index (DMI, lagged 4 months), access to hygienic flushing toilets (indoor or outdoor), access to piped water infrastructure, and the 10-year average urban land expansion rate (Table 1; Figure 2). All covariate effects were stratified by the eight subregions of Vietnam using random slopes to capture distinct patterns. The model also incorporated spatiotemporal random effects, including a monthly second-order cyclic random walk effect, a yearly independent and identically distributed effect on the dengue year (i.e. May-April) replicated by subregion, and a spatial effect with structured and unstructured components using a district-level adjacency matrix (Figure S4).

**Table 1.**
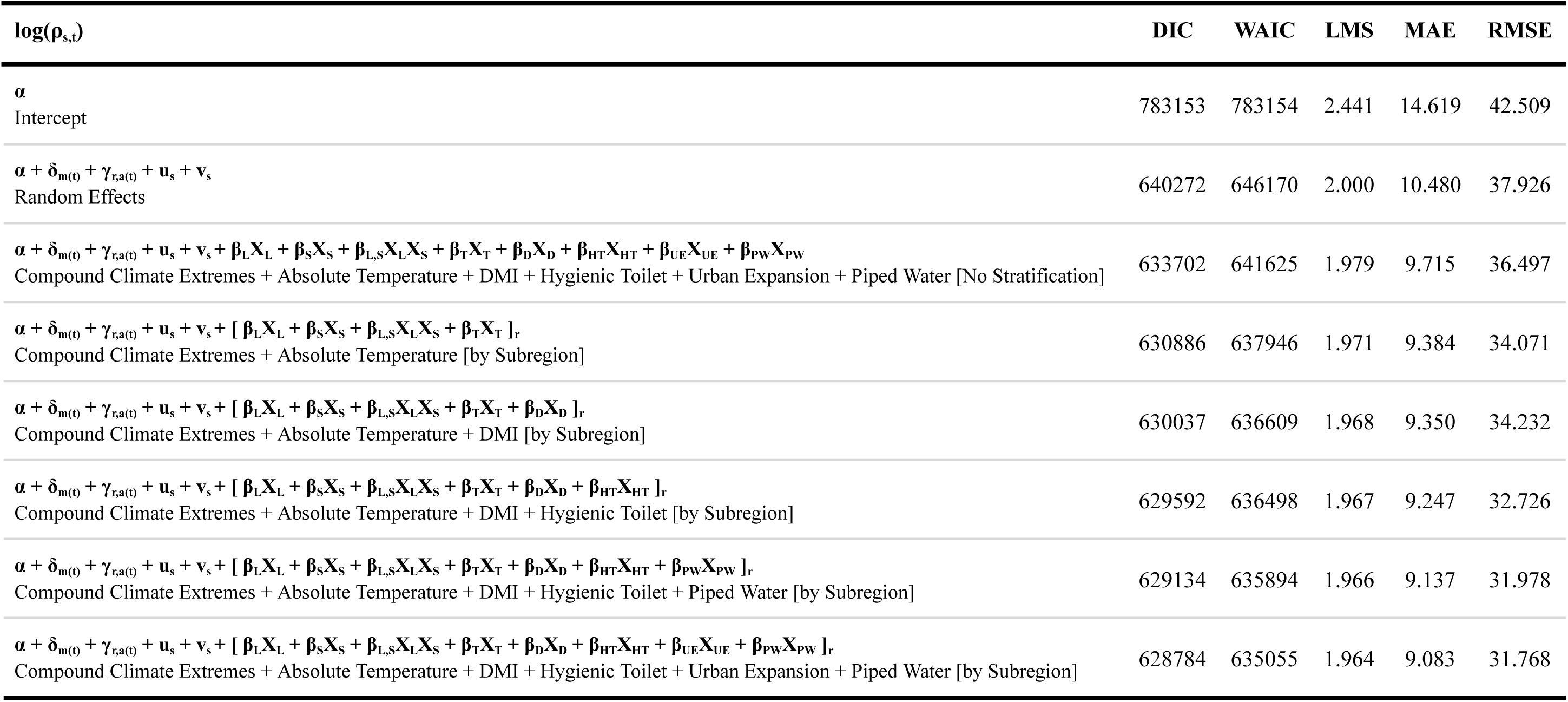
Goodness-of-fit metrics for the selected model and formulations of reduced complexity. . Models estimate the log dengue incidence rate, log(ρ_s,t_), and comprise an intercept (α), monthly random effect (δ_m(t)_), yearly random effect replicated per subregion (γr,_a(t)_), spatial random effects (u_s_ + v_s_), and fixed effects (β) applied to covariates (X) where bracketed terms […]_r_ represent stratification by subregion. Covariates are: L = SPEI-6 (lagged 5 months), S = SPEI-6 (lagged 0 months), T = 6-month absolute mean temperature (lagged 0 months), D = Dipole Mode Index (lagged 4 months), HT = hygienic toilet access, UE = 10-year urban expansion rate, PW = piped water access. Metrics are the Deviance Information Criterion (DIC), Watanabe-Akaike Information Criterion (WAIC), Log Mean Score (LMS), Mean Absolute Error (MAE), and Root Mean Squared Error (RMSE), where lower values indicate better fitting models.

**Figure 2.**
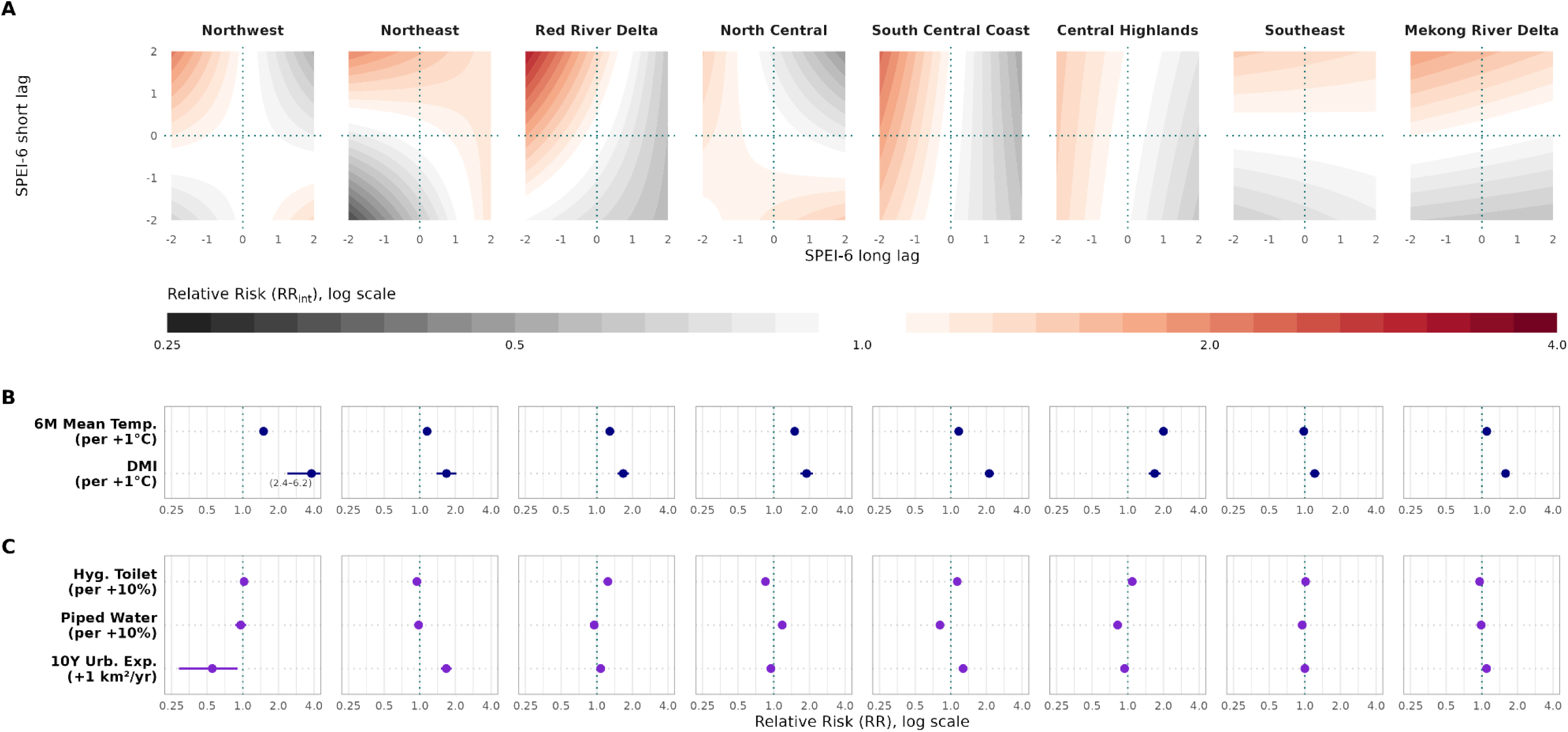
Subregionally stratified effects of compound climate extremes and additive climatic, oceanic and socioeconomic factors on dengue relative risk. Risks are estimated for the selected model with covariates stratified by eight subregions (columns). (A) Relative risk of dengue (RR_int_) for the two-way interaction between the SPEI-6 long-lag (lag 5 months) and short-lag (lag 0 months). Surfaces show the posterior mean RR_int_ over ±2 standard deviations of long-lag SPEI-6 (x-axis) and short-lag SPEI-6 (y-axis) relative to zero conditions. (B) Relative risk (RR, mean + 95% CrI) of the 6-month absolute mean temperature (lag 0 months) per +1°C, and DMI (lag 4 months) per +1°C. (C) Relative risk (RR, mean + 95% CrI) of hygienic toilet access per +10%, piped water access per +10%, and 10-year urban expansion rate per +1 km^2^/year. Credible intervals outside of the axis range are stated.

Compound dry-then-wet conditions – long-lag dry extremes followed by short-lag wet extremes – were associated with increased dengue relative risk in all eight subregions (Figure 2A). Relative risk was highest following this sequence in seven subregions (Table S1). Under dry-then-wet conditions, the risk of dengue more than doubled in the Red River Delta (RR_int_ = 2.53 [2.22, 2.83]), and South Central Coast (RR_int_ = 2.08 [1.87, 2.31]). The North Central was the exception, with peak risk increasing by 34% after wet-then-dry conditions (RR_int_ = 1.34 [1.18, 1.50]). The compound extremes associated with decreased dengue risk were more heterogeneous (Figure 2A; Table S1). Risk was lowest following persistently drier conditions in the Northwest, Northeast, and Southeast; wet-then-dry extremes in the Red River Delta, Central Highlands, and Mekong River Delta; and persistently wetter conditions across the central coastal subregions (North Central, and South Central Coast). We additionally tested a three-way interaction between these long-lag and short-lag drought indicators and the 6-month mean temperature anomaly, which modified the dry-then-wet sequence along a latitudinal gradient, favouring cooler anomalies at higher latitudes and warmer anomalies at lower latitudes except the Mekong River Delta (Figure S5). However, the three-way interaction did not consistently improve model goodness-of-fit.

Higher 6-month mean temperatures were associated with increased dengue risk across all North and Central subregions (Figure 2B). Per 1°C increase, the association was strongest in the Central Highlands (RR = 2.00 [1.93-2.07]), and weakest in the Mekong River Delta (RR = 1.10 [1.06, 1.14]), with no association evident in the Southeast (RR = 0.98 [0.94, 1.02]). A 1°C increase in the Dipole Mode Index, the sea surface temperature gradient between the western and southeastern tropical Indian Ocean, was associated with increased risk across all eight subregions (Figure 2B). This association was strongest in the Northwest with high uncertainty (RR = 3.81 [2.37, 6.18]), and weakest in the Southeast (RR = 1.21 [1.10, 1.33]).

Socioeconomic effects were heterogeneous across subregions in both direction and magnitude (Figure 2C). A 10% increase in hygienic toilet access was associated with greater dengue risk in three of the four most urbanised subregions (Red River Delta, South Central Coast, and Central Highlands), and lower risk in three of the four least urbanised (Northeast, North Central, and Mekong River Delta). Piped water access reversed this pattern, with lower risk associations across the four most urbanised subregions, and greater risk only in the North Central. Urban land expansion associations were more mixed. Per 1 km^2^/year increase, the rate was associated with increased dengue risk in the Northeast, Red River Delta, South Central Coast, and Mekong River Delta, and with decreased risk in the Northwest, North Central, and Central Highlands. Iteratively removing these covariates consistently degraded goodness-of-fit metrics (Table 1; Figure S6).

### Human mobility influences dengue risk along an urban-rural gradient

Using movement range maps from Meta AI for Good (Meta, 2024), we derived three covariates representing mobility across Vietnam (Figure 3A-C). Staying probability is the probability that a resident remains within their home district, with higher values indicating a less mobile population. Conditional outgoing entropy describes the geographical spread of people leaving a district, with higher values indicating dispersed movement to many destinations. Inflow quantifies the volume of arrivals from other districts, with higher values indicating greater mobility into the district. Human movement was largely concentrated around major urban districts, such as Hanoi and Ho Chi Minh City.

**Figure 3.**
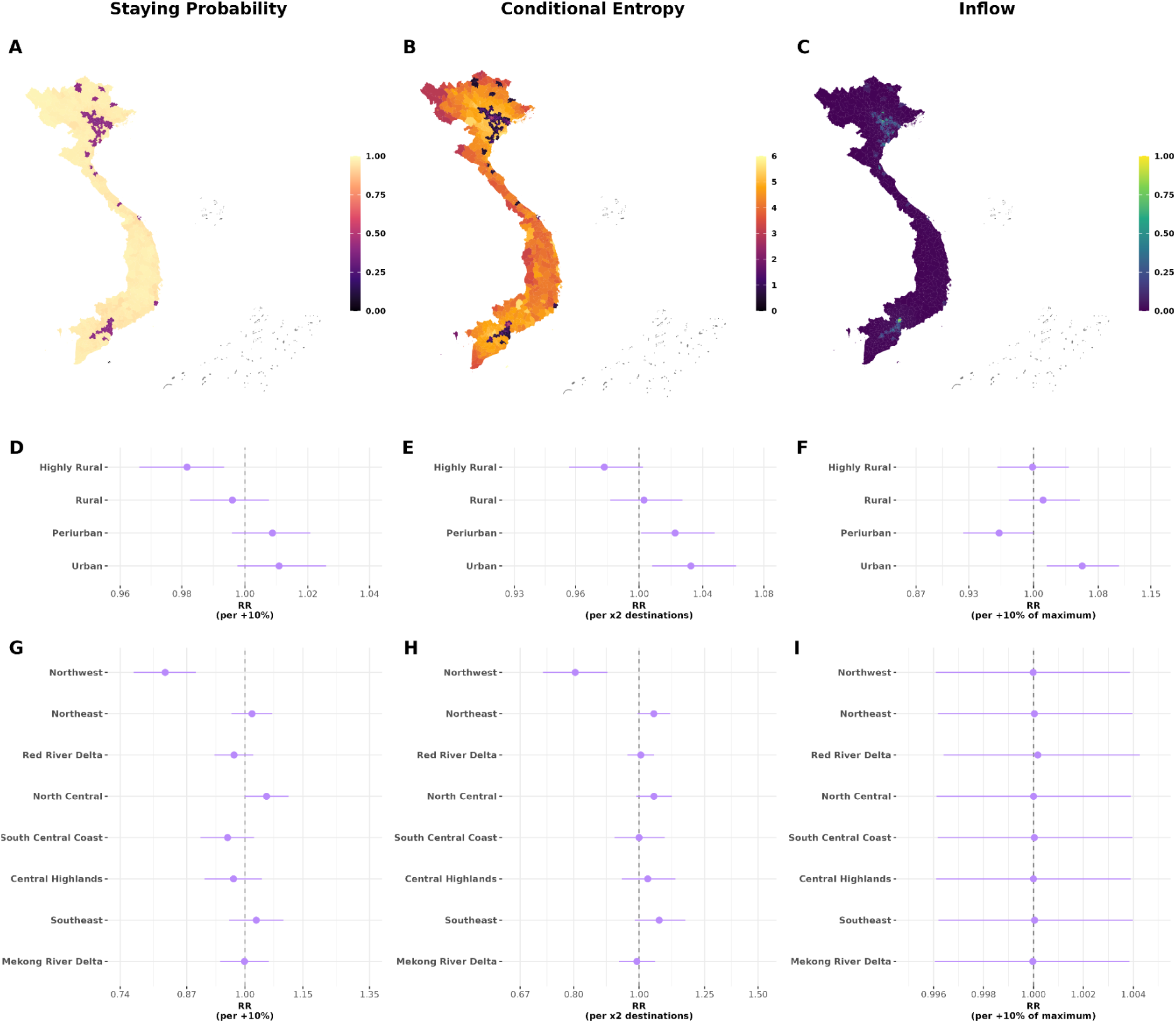
Human mobility association with dengue relative risk, stratified by urbanisation level and subregion. (A-C) District-level distributions of three mobility covariates derived from Meta AI for Good movement range maps: (A) staying probability, (B) conditional outgoing entropy, and (C) relative inflow. (D-F) Relative risk (RR, mean + 95% CrI) of each mobility covariate on dengue incidence stratified by urbanisation level (Highly Rural, Rural, Periurban, Urban): (D) staying probability per 10% increase, (E) conditional outgoing entropy per doubling of destinations, and (F) relative inflow per 10% increase of the maximum. (G-I) Relative risk (RR, mean + 95% CrI) of each mobility covariate on dengue incidence stratified by subregion: (G) staying probability per 10% increase, (H) conditional outgoing entropy per doubling of destinations, and (I) relative inflow per 10% increase of the maximum. Mobility associations were estimated in the best-fitting model which included covariate effects stratified by eight subregions for SPEI-6 (lagged 5 months) interacted with SPEI-6 (lagged 0 months) and additive 6-month absolute mean temperature (lagged 0 months), DMI (lagged 4 months), hygienic toilet access, piped water access, and 10-year urban expansion rate.

We added each covariate individually to the best-fitting socio-climatic model, testing linear, nonlinear and spatially stratified effects. Stratification by urbanisation level or subregion marginally improved some goodness-of-fit metrics (Figure S6). Mobility covariates stratified by urbanisation showed a gradient in the dengue relative risk from the most rural to the most urban districts (Figure 3D-F). In highly rural districts, dengue risk was highest where residents left their district more often and where movement was concentrated to fewer destinations. This pattern reversed with increasing urbanisation. In urban districts, risk was highest where residents were more likely to remain in their district and where outgoing travel was most widely dispersed. Furthermore, dengue risk increased in urban districts with greater volumes of arrivals. Stratification by subregion (Figure 3G-I) revealed the same rural pattern in the Northwest, the most rural subregion, where decreased staying probability and conditional entropy increased dengue risk.

We substituted the adjacency matrix in the spatial random effect with alternative movement matrices, including gravity models, travel time estimates, and human mobility probabilities between districts, to define spatial dependence by connectivity. No movement matrix improved all goodness-of-fit metrics relative to adjacency. The combined structured and unstructured components of the spatial random effect were consistent in magnitude across all specifications, whereas the structured component was lower for each movement matrix compared to adjacency (Figure S7).

### Spatially stratified climatic and socioeconomic effects improve dengue predictability

We compared results from rolling-origin cross-validation, where each model is refitted at successive time points with historical data to predict the following 1-6 months, for five model formulations (Table S2). M_1_ represents the best-fitting model discussed above, which incorporates climatic and socioeconomic covariates stratified by eight subregions along with spatial, monthly, and yearly random effects. M_2_ excludes the yearly random effect from the best-fitting model, and M_3_ estimates covariate effects nationally without subregional stratification. B_1_ and B_2_ are the corresponding baseline models, which retain the random-effects structures of M_1_ and M_2_ respectively but exclude all covariates. Predictive skill, pooled across all districts-months, was compared across lead times of 1 to 6 months using the Continuous Ranked Probability Score (CRPS) for case estimation, Brier Score (BS) for the accuracy of probabilistic predictions, and the Area Under the ROC Curve (AUC) for outbreak detection, with the associated True Positive Rate (TPR; sensitivity) and False Positive Rate (FPR; 1-specificity) (Figure 4; Table S3). A lower CRPS and BS, and higher AUC, indicate better predictive performance.

**Figure 4.**
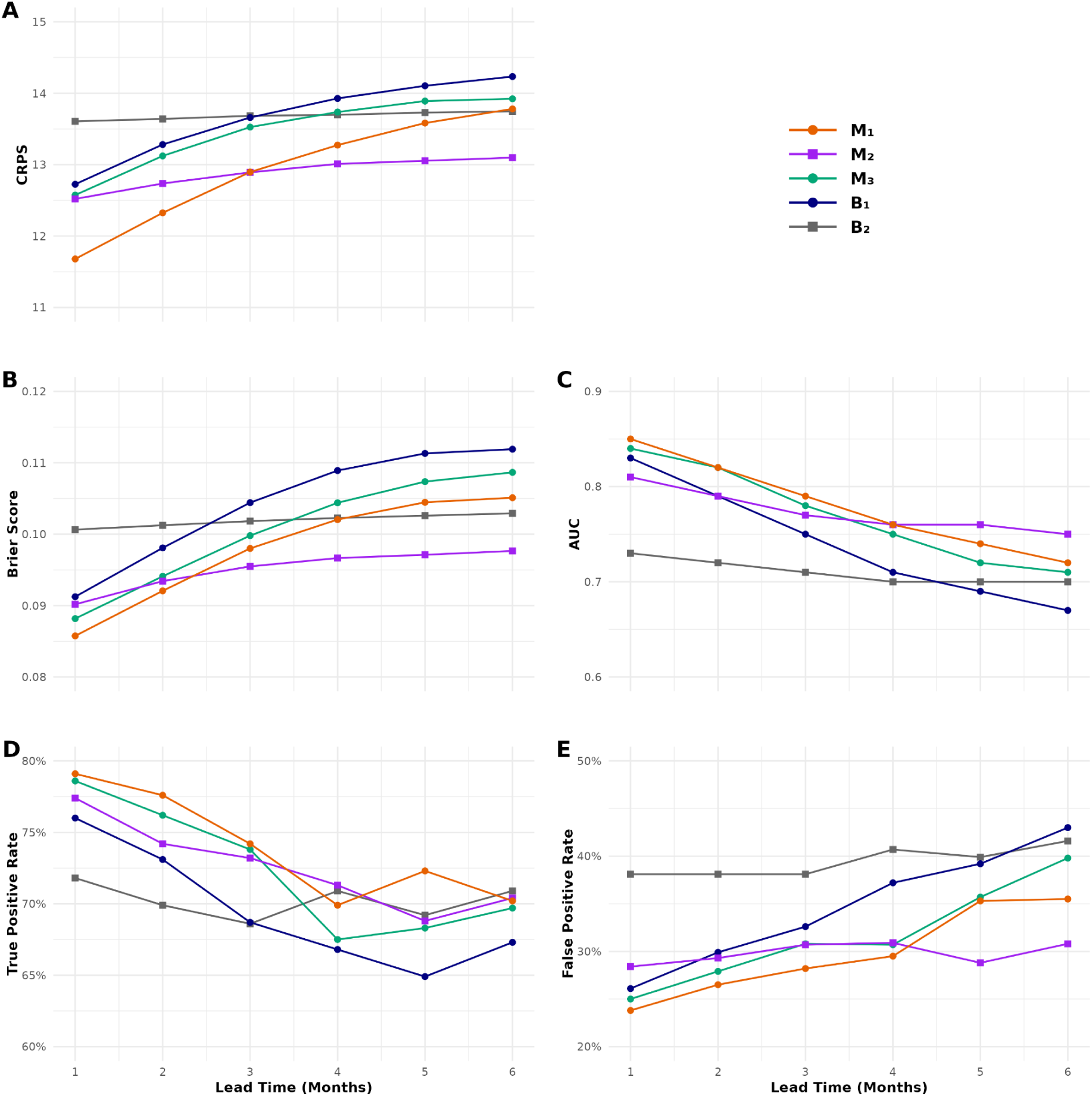
Predictive skill over all district-months for five model formulations at lead times of 1 to 6 months. Models were evaluated by rolling-origin cross-validation over May 2016 to April 2021. Panels show (A) Continuous Ranked Probability Score (CRPS) for case estimation, (B) Brier Score for probabilistic accuracy, (C) Area Under the Receiver Operating Characteristic Curve (AUC) for outbreak detection, (D) True Positive Rate, and (E) False Positive Rate. Lower values indicate better performance for the CRPS, Brier Score and the False Positive Rate, whereas higher values indicate better performance for the AUC and True Positive Rate. M_1_ represents the best-fitting model with socio-climatic covariates stratified by subregion and spatial, monthly, and yearly random effects. M_2_ omits the yearly random effect, and M_3_ estimates covariate effects nationally without subregional stratification. B_1_ and B_2_ are baseline models with no covariates but corresponding random-effects structures of M_1_ and M_2_ respectively.

Incorporating socio-climatic covariates improved case estimation, probabilistic accuracy and outbreak detection at all lead times of 1 to 6 months, comparing the best-fitting model (M_1_) with its baseline (B_1_). These results were consistent when we excluded the yearly random effect from the models (M_2_ vs B_2_). By allowing covariate effects to vary by subregion (M_1_), predictive skill improved across all metrics and lead times compared to unstratified effects (M_3_).

Comparing baseline models, we found that including the yearly random effect, which accounts for interannual variation in dengue incidence, improved case estimation by up to 6.5%, probabilistic accuracy by up to 9.3%, and outbreak detection by up to 0.1 unit in AUC at shorter lead times, but worsened predictive skill as lead time increased. When comparing socio-climatic models, we found similar short-lead improvements with the yearly random effect included, and worsening of predictive skill from lead times of 3, 4 or 5 months depending on the metric.

### Predictive skill is highly variable across spatial scales and lead times

We evaluated predictive skill by subregion and district across 1-6-month lead times for the two stratified socio-climatic formulations, M_1_ and M_2_, and their corresponding baselines, B_1_ and B_2_ (Figure 5; Table S4-S6). At least one socio-climatic model improved case estimation, probabilistic accuracy and outbreak detection relative to its baseline at every lead time in five of eight subregions, and across most metrics and leads in two further subregions. The Northeast was the exception, where improvement was restricted to M_2_ at the 1-month lead. District-level performance showed greater variability (Figure 5B-C). Either socio-climatic formulation was the best-performing model in 58.9-63.3% of districts for case estimation and 52.5-55.6% for probabilistic accuracy across lead times, rising to 66.0-71.4% and 59.8-70.1% respectively in the four subregions with the highest dengue incidence.

**Figure 5.**
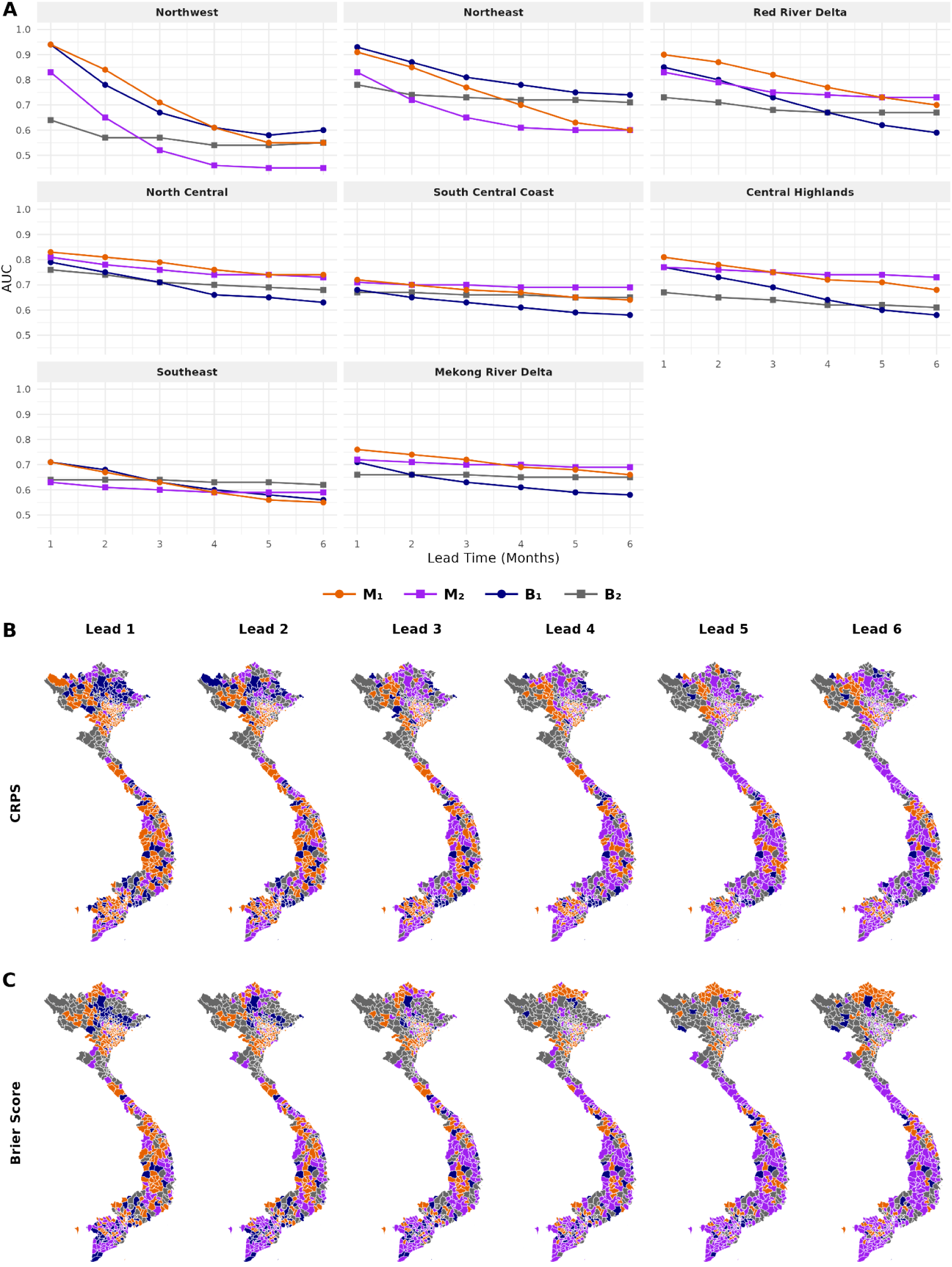
Subregional and district-level predictive skill of models M_1_, M_2_, B_1_, and B_2_ across lead times of 1 to 6 months. (A) AUC by subregion and lead time for the four models. (B-C) Best-performing model per district by (B) CRPS and (C) Brier Score, at each lead time (columns) coloured by model. Maps only show the 670 districts modelled.

Inclusion of the yearly random effect (M_1_) improved outbreak detection across lead times at higher latitudes compared to its exclusion (M_2_), but only for shorter leads at lower latitudes (Figure 5A; Table S6). Case estimation and probabilistic accuracy similarly showed short-lead improvements at lower latitudes with the yearly random effect included, but largely degraded skill across lead times at higher latitudes, except the Northwest (Figure 5B-C; Table S6). No single formulation was optimal across all locations, metrics, and lead times.

We evaluated subregion-specific outbreak detection skill alongside the TPR and FPR at operational lead times of 1 to 3 months (Figure S8). We defined adequate performance as capturing at least two-thirds of outbreak months (TPR ≥ 67%) whilst falsely alerting no more than one-third of non-outbreak months (FPR ≤ 33%). Six of eight subregions met both criteria at 1- and 2-month leads (Northwest, Northeast, Red River Delta, North Central, Central Highlands, and Mekong River Delta), with AUC values of 0.74-0.94. Only four subregions remained adequate at the 3-month lead. The South Central Coast and Southeast did not meet both criteria at any lead time, with false alarms ranging 37-43% and AUC values of 0.63-0.72.

## Discussion

Here, we demonstrate that compound climate extremes, socioeconomic conditions, and human mobility shape dengue dynamics heterogeneously across Vietnam. Using 20 years of data, we developed a Bayesian modelling framework to quantify associations with dengue relative risk at the district level, stratifying covariate effects by the country’s eight subregions. Interacting long-lag and short-lag drought indicators, we show that dengue relative risk peaked following compound dry-then-wet conditions in seven subregions. Higher mean temperatures increased risk across North and Central Vietnam, but showed weak-to-null effects in the South. Stratifying mobility effects by urbanisation revealed an urban-rural gradient, where dengue risk increased with greater outward movement to fewer destinations in highly rural districts, and with population retention and increased arrivals into urban districts. Incorporating key climatic and socioeconomic covariates, we evaluated model predictive skill at lead times of 1 to 6 months, operationally relevant for early warning in Vietnam. Subregional stratification of climatic and socioeconomic covariates improved dengue forecasting across all lead times relative to unstratified and baseline models, although predictive skill varied substantially across spatial scales.

Previous compound event analyses of dengue have explored the effects of concurrent extremes pooled across diverse settings (D. Wang et al., 2026; Y. Wang et al., 2024). In this study, we quantify the effects of sequential extremes on dengue risk by interacting the SPEI-6 at two lags, with effects stratified by subregion. Dengue relative risk increased after compound dry-then-wet conditions in all eight subregions, and showed the maximum risk in seven subregions, consistent with findings in the Caribbean (Fletcher et al., 2025). Within the compound event typology, this sequence may represent a preconditioned event, where antecedent dry extremes create conditions that amplify the effect of subsequent wet extremes, or a temporally compound event, where drier and wetter conditions each contribute to transmission such that successive events produce a greater joint impact (Zscheischler et al., 2020). During drier conditions, reduced access to piped water can drive households to store water in domestic containers that are productive *Aedes* habitats (Gómez-Vargas et al., 2024; Stewart-Ibarra et al., 2013). In Vietnam, reduced water supply has been associated with a higher rate of dengue hospitalisation (Schmidt et al., 2011). Containers are hypothesised to become less actively managed once drought eases, providing abundant larval habitats as wetter conditions return (Lowe et al., 2018). Under a preconditioned interpretation, drought-related storage would supply the habitat that allows subsequent wet conditions to drive risk. Under a temporally compounding interpretation, both conditions would represent distinct pathways that lead to cumulative effects. A previous district-level study in Vietnam found that increased piped water access mitigated dengue risk associated with moderately dry conditions several months prior, consistent with storage-driven transmission (Gibb et al., 2023). However, installing household taps and tanks did not reduce water storage or *Aedes* abundance in the Mekong Delta, with households favouring stored rainwater due to cost, quality and supply reliability (Tran et al., 2012). Piped water is therefore an imperfect marker of storage behaviour as cultural preferences and supply stability may dictate local practices. We consequently integrated piped water access as an independent variable stratified by subregion to flexibly capture localised dynamics.

The absolute 6-month mean temperature was associated with increased dengue risk across all North and Central subregions, particularly at higher latitudes and altitudes. A 1°C temperature rise increased dengue relative risk by 100% in the Central Highlands, 50% in the Northwest and North Central, and 29% in the Red River Delta. Associations were weak-to-null across the South, where consistently warm conditions vary within a narrow thermal range. These patterns may be partly explained by mosquito thermal biology. Trait-based models predict thermal optima for dengue transmission at 26.4°C for *Aedes albopictus*, and 29.1°C for *Aedes aegypti* (Mordecai et al., 2019). Previous district-level analysis in Vietnam identified an unstratified national unimodal temperature-dengue relationship, with risk peaking around 27.0°C (Gibb et al., 2023). Temperatures across North and Central Vietnam were typically below these optima, so warming brings conditions closer to those favouring transmission. Conversely, mean temperatures in the South were 26.9°C in both subregions with narrow interquartile ranges spanning 0.9-1.1°C, indicating favourable conditions across most of the year. *Aedes* distribution in Vietnam follows a latitudinal gradient, where sampling across 18 urban sites found *Aedes albopictus* predominant in the North, and *Aedes aegypti* in the South (Duong et al., 2022). *Aedes aegypti* is regarded as the primary dengue vector, with limited evidence on the transmission contribution of *Aedes albopictus* across Vietnam (Kim Lien et al., 2015; Whitehorn et al., 2015). *Aedes aegypti* populations may also differ in thermal response, with extrinsic incubation temperature affecting dengue virus infection rates differently in mosquitoes from Hanoi compared to Ho Chi Minh City (Gloria-Soria et al., 2017).

The Indian Ocean Dipole acts upon dengue indirectly by modulating local climate (Chen et al., 2024), with reported associations across South and Southeast Asia inconsistent in direction. For example, a higher DMI increased dengue relative risk in Bangladesh (Banu et al., 2015), but decreased cumulative relative risk in Lao PDR (Houatthongkham et al., 2026), and showed no association across Indonesia (Djaafara et al., 2026). In Southern Taiwan, the DMI tracked interannual dengue patterns but acted more strongly on local climate (Chuang et al., 2017). Here, a positive DMI lagged 4 months was associated with increased dengue relative risk across all eight subregions. These findings align with evidence from coastal provinces in Central Vietnam, where the DMI was positively associated with dengue incidence (L. T. Nguyen et al., 2020). However, the climatic pathway potentially underpinning this relationship is unclear. A positive DMI has been linked to excess precipitation across mainland Southeast Asia (Phan-Van et al., 2022), yet the same phase has been associated with drier autumn conditions across the region (Nguyen-Le et al., 2024), and reduced annual maximum precipitation in Vietnam (Do et al., 2020). Further work characterising local teleconnections and its relationship with dengue would aid interpretability.

The association between mobility and dengue varied along the urban-rural gradient. In highly rural districts, higher dengue risk was found with greater outgoing mobility to fewer destinations, whereas in urban districts, higher dengue risk was found with greater incoming mobility and more dispersed outgoing mobility. These findings are consistent with connectivity to urban centres amplifying dengue transmission in susceptible populations (Harish et al., 2024; Salje et al., 2021). Stratification by subregion reproduced the rural pattern in the Northwest, partially aligning with the hypothesis that mobility exerts greater influence across the subtropical North than the endemic South (Gibb et al., 2023). Despite this, mobility did not improve predictive skill in the Northwest, nor consistently across metrics in other subregions (Figure S9-S11). Furthermore, replacing the adjacency matrix with alternative movement matrices did not improve model fit, indicating that residual spatial variation in dengue risk was adequately captured by district-level contiguity. Due to limited availability, mobility data were collected outside of our study period in 2023-2024, and were assumed static over time, representing a major limitation in this study. Long-term temporally-resolved data could model mobility more dynamically, capturing infrastructural changes, seasonal travel, or behaviour during mass events.

Spatially stratifying covariate effects via random slopes substantially improved model fit and predictive skill across 1-6-month lead times. This stratification likely captures complexity in dengue dynamics which unstratified effects may obscure, such as differences in standing population immunity, epidemiology, climatology, vector distribution and infrastructure. Stratification by subregion improved fit more than by other spatial classifications, such as elevation, urbanisation, and Köppen-Geiger climatic zones, possibly because subregions capture these aspects simultaneously. Subregionally stratified climatic and socioeconomic covariates improved predictive skill at every lead time compared to national unstratified estimates and baseline models. However, predictive improvements were spatially heterogeneous. Outbreak detection improved at most operational lead times in six of eight subregions. District-level case estimation and probabilistic accuracy gains were more modest, although were greatest in areas with the highest dengue incidence. These improvements demonstrate that climatic and socioeconomic conditions add predictive value beyond the variation captured by random effects alone.

Outbreak detection is central to the operational utility of early warning systems, providing actionable information to inform public health preparedness and response, such as procuring clinical supplies, conducting community outreach, and deploying targeted vector control (Alcayna et al., 2025; Díaz et al., 2024; Fletcher et al., 2025). Operational trigger thresholds could be calibrated to prioritise true detection or reduce false alerts, which would require assessment of the local response capacity, decision-maker needs, and relative costs of missed outbreaks against unnecessary mobilisation. For the outbreak threshold, we selected the mean plus two standard deviations, previously identified as the optimal province-level threshold in Vietnam (Colón-González et al., 2021). Thresholds inherently identify outbreaks with differing frequency, duration and case burden across settings (Brady et al., 2015), therefore AUC comparisons within a single subregion are likely to be more robust than comparisons across subregions. Outbreak detection was weakest in the Southeast and South Central Coast. Due to historically high incidence, epidemics arise due to a complex nexus of immunity, circulating serotypes, vector control activities, and human and healthcare-seeking behaviour (Brook et al., 2024; Finch et al., 2025; García-Carreras et al., 2022), among which the influence of climate is likely more limited. Extending our framework to incorporate population susceptibility and interventions, where such data are available, may enhance predictive skill in settings where transmission is sustained year-round.

The yearly random effect introduced a trade-off that shifted with lead time. Its inclusion improved predictive skill at shorter leads, but eventually degraded performance as lead time increased. Subregional outbreak detection showed similar results at lower latitudes, but inclusion of the yearly effect improved outbreak detection across most leads at higher latitudes. This is consistent with Vietnam’s distinct epidemiological profiles, where interannual variation is large relative to seasonal variation in the emerging North, whereas transmission is sustained and strongly seasonal toward the South (Colón-González et al., 2021). We fixed the dengue season from May to April during cross-validation, meaning the number of within-season months informing the yearly random effect varied systematically with prediction month, potentially confounding skill with seasonality. We therefore compared our cross-validation scheme against a rolling season definition, which anchored the number of within-season months to be constant across the year (Text S1; Figure S12-S16). The fixed definition improved case estimation and probabilistic accuracy in 63-72% of districts across lead times, and outbreak detection at lead times of 1-3 months. Improvements were further demonstrated during the peak season, from May to November.

Some limitations warrant consideration. We used observed climate covariates during cross-validation to attribute predictive skill to the modelling framework instead of the climate forecasts that would drive the system operationally. Seasonal climate forecasts can extend the potential warning horizon of dengue by up to 6 months, although introduce additional uncertainty that would propagate through the modelling framework, with skill varying by covariate, region, season, and lead time (Wanthanaporn et al., 2025; Weisheimer & Palmer, 2014). Vietnam’s recent administrative reform removed district-level units, meaning the framework represents a proof-of-concept rather than a deployable system. Given recurrent boundary changes, geolocated point-based frameworks would allow models to remain independent of shifting borders yet aggregable to decision-relevant units, representing an avenue for future work. Our case definition included both confirmed and suspected cases which may incorporate co-circulating arboviruses, such as chikungunya or Zika virus, although reported activity of both diseases was low relative to dengue over this period (C. T. Nguyen et al., 2020; Quan et al., 2018). We also did not directly account for interventions that alter transmission, including *Wolbachia* deployments (Hien et al., 2022; T. H. Nguyen et al., 2015) or dengue vaccination (Sáez-Llorens et al., 2025), which may become more important in the future if widely applied. Despite these limitations, our findings show that compound climate extremes, socioeconomic conditions, and human mobility act heterogeneously across Vietnam, and that accommodating these differences enhances the predictability of dengue incidence and outbreaks. These advances support the development of tailored fine-scale early warning in Vietnam, and could be adapted to enhance modelling approaches in other settings with spatially varying climate-sensitive disease risk.

## Methods

### Study area and dengue data

Vietnam is located in Southeast Asia, with a land area of around 331,000 km^2^ (Worldometer, 2026a), consisting of three regions (North, Central, and South) and eight subregions (Northeast, Northwest, Red River Delta, North Central, South Central Coast, Central Highlands, Southeast, and Mekong River Delta) (Figure 1). Vietnam is the 16th most-populous country globally with a population of over 102 million inhabitants (Worldometer, 2026b).

Dengue surveillance data were obtained from the Pasteur Institute Ho Chi Minh City, Pasteur Institute Nha Trang, Tay Nguyen Institute of Hygiene and Epidemiology, and National Institute of Hygiene and Epidemiology, processed for previous research in Vietnam (Gibb et al., 2023). Confirmed and suspected cases recorded in the passive surveillance system from May 2001 to April 2021 were included. Dengue cases were aggregated temporally at the monthly level and spatially at the district level. Due to changes in district-level administrative boundaries over the study period, some districts were merged to align with the Global Administrative Areas (GADM) boundaries for data interoperability (GADM, 2026), where a total of 670 were included. In July 2025, administrative boundaries in Vietnam were reformed such that 63 provinces and cities were merged into 34 first-level subdivisions, district-level units were removed, and 10,035 commune-level units were merged into 3321 second-level subdivisions (VietNamNet Global, 2025). Despite this reform, district-level aggregation was retained in this study to reflect dengue reporting from 2001 to 2021 and serves as a proof of concept for fine-scale predictive modelling.

### Climate and environmental data

We collated temperature and precipitation data from the ERA5-Land global reanalysis dataset (0.1° x 0.1°) across our study area from January 1981 to December 2021. For eight districts, we alternatively sourced data from the ERA5 reanalysis dataset (0.25° x 0.25°) as these were small islands or coastal areas not covered by ERA5-Land. Gridded data were spatially mean-aggregated to the district level using the exactextractr package in R (Baston et al., 2025). We derived temperature and precipitation indices for 1-, 3-, 6- and 12-month rolling windows lagged by 0-6 months. Temperature indices included mean, minimum and maximum absolute temperature (°C) and temperature anomalies (°C), defined as the difference between the observed temperature and location-month-specific climatology for the reference period 1991-2020. Precipitation or drought indicators included total precipitation (mm), the Standardised Precipitation Index (SPI), and the Standardised Precipitation Evapotranspiration Index (SPEI). SPI and SPEI were calculated using gamma and log-logistic distributions, respectively, compared to the location-month-specific baseline from 1980-2020. SPI quantifies rainfall anomalies solely using precipitation data, whereas SPEI estimates the water balance by accounting for potential evapotranspiration using both precipitation and temperature data. For both drought indicators, negative or positive values indicate drier or wetter conditions, respectively, than the historical baseline.

Oceanic indices, including the Dipole Mode Index (DMI) and the Oceanic Niño Index (ONI), were collected from the National Oceanic and Atmospheric Administration (NOAA) website (https://psl.noaa.gov/data), and lagged by 0-6 months. DMI data are obtained from the HadISST1.1 dataset. DMI describes the intensity of the Indian Ocean Dipole (°C), representing the anomalous sea surface temperature (SST) gradient between the western equatorial Indian Ocean (50E-70E, 10S-10N) and the southeastern equatorial Indian Ocean (90E-110E, 10S-0N). A positive DMI represents warmer-than-normal waters toward the west and cooler-than-normal waters toward the southeast equator, whereas a negative DMI represents the inverse. ONI data are obtained from the ERSSTv5 dataset. ONI represents the centred 3-month running mean SST anomalies in the Niño 3.4 region of the tropical Pacific Ocean (5N-5S, 120W-170W). A positive ONI reflects warming, where +0.5°C indicates an El Niño, and a negative ONI reflects cooling, where a -0.5°C indicates a La Niña.

High-resolution (1 km) Köppen-Geiger climate classification data were obtained for the 1991-2020 reference period (Beck et al., 2023). Each district was assigned a climate classification by calculating the mode across all grid cells within the polygon. We used the Copernicus GLO-90 Digital Elevation Model (European Space Agency, 2024), available at the 90 m grid level, to calculate the median elevation per district. We then classified districts into elevation bands by either 2, 3, 4, 5, 6 or 8 groups.

### Socioeconomic and demographic data

Here, we considered only socioeconomic variables previously collated and identified as key predictors of dengue at the district level in Vietnam (Gibb et al., 2023). These variables included the proportion of district households with access to hygienic flushing toilets and access to piped water infrastructure, estimated via interpolation between the Vietnam Population and Housing Censuses (National Statistics Office of Vietnam, 2010, 2020). Additionally, the 10-year urban land expansion rate, derived from Landsat 30 m annual impervious surface data (Liu et al., 2020), was included. District-level total and urban population figures were obtained from census data and linearly interpolated to estimate yearly totals. The annual population per district was used to calculate the monthly dengue incidence rate per 100,000 inhabitants. We assigned each district an annual urbanisation level using quartiles from the proportion of inhabitants living in urban areas across our dataset, defined as Highly Rural ([0.0, 5.0]%), Rural ([5.1, 10.5]%), Periurban ([10.6, 52.9]%), and Urban ([53.0, 100.0]%). Over the 20-year period, 503 districts held the same urban classification, whereas 129 districts changed classification once, 27 districts changed twice, and 11 districts changed three times (Figure S1).

### Mobility data

We derived mobility matrices and covariates from multiple sources, including demographic data, travel time estimates between districts (Gibb et al., 2023), and Meta AI For Good (Meta, 2024). For mobility matrices, we included distance-decay gravity models, travel time estimates, and human mobility probabilities between all pairwise combinations of districts (Text S2). Gravity models assume that human mobility is solely driven by both population size, estimated here for 2021, and distance, calculated between centroids. Travel time represents the population-weighted mean travel time from the centroid of one district to all grid cells in another district (Gibb et al., 2023). Human mobility probabilities between districts were estimated from direct measurements of social media and call data records collected between 1 June 2023 to 1 June 2024 and transformed into movement range maps that were available via Meta AI For Good (Meta, 2024). Unlike other sources, these data additionally account for the proportion of individuals who remain within their home district, defined as the staying probability. All matrices were symmetrised, assuming that mobility between two districts is bidirectional and equal in magnitude. By default, the diagonal elements of these matrices, representing within-district mobility, were set to zero. However, we also explored alternative representations of the diagonal. For both the gravity model and travel time, we tested the integration of staying probability from the human mobility probability matrix. For the travel time, we additionally tested the mean travel time from the district centroid to all grid cells in that same district.

For mobility covariates, we included staying probability, conditional outgoing entropy, and relative inflow per district, estimated from the Meta AI for Good movement range maps (Text S2). Staying probability represents the proportion of individuals who remain in their home district, where 0 indicates total mobility and 1 indicates no mobility. Conditional outgoing entropy describes the geographical spread of people leaving a district, where lower values indicate concentrated movement to fewer destinations and higher values indicate greater dispersal to many locations. Inflow quantifies the relative volume of individuals arriving into a district from all other origins, normalised from 0 to 1, where lower values indicate less inflow and higher values indicate greater inflow. Mobility covariates were well-correlated, with absolute magnitudes ranging from 0.51 to 0.89 (Figure S17).

### Model specification

We formulated a Bayesian hierarchical mixed model using dengue case counts, y_s,t_, per district, s (1 to 670), per month, t (1 to 240), as the response variable. These counts were assumed to follow a negative binomial distribution, accounting for potential overdispersion, such that:

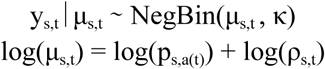

where μ_s,t_ represents the expected number of dengue cases, and κ is the overdispersion parameter, which accounts for variance exceeding the mean. Within the log-link function, p_s,a(t)_ represents the annual district-level population offset (per 100,000 inhabitants), allowing the model to adjust for yearly demographic changes, whilst ρ_s,t_ represents the expected dengue incidence rate for a given district-month.

We first defined a random-effects framework to capture the underlying spatiotemporal dependence and serve as a baseline model. This formulation included a global intercept, α, alongside temporal and spatial random effects. A dengue season was defined from May to April the following year. To account for seasonal variation, we included a temporal random effect, δ_m(t)_, for each month, m(t), from 1 to 12, specified as a second-order cyclic random walk model. Interannual variation was captured by a temporal random effect, γ_sr(s),a(t)_, replicated by subregion, sr(s), for each dengue year, a(t), from 1 to 20, specified as an independent and identically distributed variable. Finally, spatial variation was captured using structured and unstructured spatial components, u_s_ + v_s_, for each district, s, from 1 to 670, specified as a modified Besag-York Mollié (BYM2) model. The structured component is modelled such that its spatial dependence is defined by a spatial connectivity matrix, which defaults to an adjacency matrix for the baseline model. The baseline model was therefore formulated as:

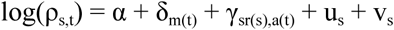

To evaluate the effects of climate, socioeconomic conditions, and human mobility, we expanded the baseline model to incorporate fixed effects, including a two-way interaction between long-lag (3-6 months) and short-lag (0-3 months) drought indicators to capture compound climate extremes, as well as additive covariates. Random slopes enable covariate parameter estimations to be spatially stratified, with a separate effect per stratum. We incorporated random slopes to account for spatially-varying dengue dynamics, testing stratifications by region, subregion, Köppen-Geiger climate classification, urbanisation level, or elevation bands with 2-8 groups. The expanded model structure is formulated such that:

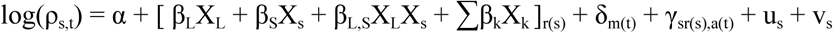

where X_L_ and X_s_ represent the long-lag and short-lag drought indicators, respectively, for a given district-month, and X_k_ other additive covariates including climatic, socioeconomic and mobility variables, with coefficients, β, that correspond to individual or interactive effects. The bracketed notation denotes that the coefficients are estimated as random slopes by the spatial stratification, r(s), assigned to the district, s.

Model parameters were estimated in R version 4.6.1 using the Integrated Nested Laplace Approximation (INLA) version 23.04.24 (Rue et al., 2009) implemented via the package GHRmodel version 0.1.1 (Milà et al., 2025).

### Model development and selection

We employed a stepwise model development procedure in which the best-performing combinations of lagged indices at each stage were carried forward (Figure S3). Candidate models were evaluated using five goodness-of-fit metrics, where lower values indicate improved performance. The Deviance Information Criterion (DIC) assesses the trade-off between model fit and complexity based on a point estimate of the posterior parameters. The Watanabe-Akaike Information Criterion (WAIC) provides a fully Bayesian evaluation of the posterior distribution which offers a robust penalty for overfitting. The Log Mean Score (LMS) evaluates the model’s out-of-sample predictive calibration and sharpness via a leave-one-out cross-validation approach. The Mean Absolute Error (MAE) quantifies the average absolute deviation between modelled and observed cases. The Root Mean Squared Error (RMSE) calculates the square root of the average squared errors, penalising models that fail to capture larger peaks in cases. Furthermore, we favoured models demonstrating credible effects defined as the 95% credible interval of the effect excluding zero, and reduced random-effect variance compared to the baseline model (Fletcher et al., 2025).

First, we established the baseline model with appropriate spatiotemporal random effects. Climate covariates were then integrated iteratively to minimise collinearity, starting with an univariable analysis, followed by bivariable and multivariable additive models, and then interaction models. We subsequently integrated top-performing oceanic and socioeconomic covariates into the best candidate climatic models. Finally, mobility was evaluated through two distinct mechanisms - either by incorporating standalone mobility covariates, or by substituting the spatial adjacency matrix with a movement matrix. The most robust framework emerging from this final stage was selected to assess predictive performance via cross-validation.

### Cross-validation

We evaluated model predictive skill using rolling-origin cross-validation with an expanding window on five years of observations from May 2016 to April 2021 at lead times of 1 to 6 months. Models were trained on historical data from May 2001 up to *n* months before the prediction month for lead time *n*, with the training window expanding by one month for each successive prediction. Probabilistic forecasts were generated by drawing 500 samples from the posterior distribution. We assumed that dengue forecasts were issued on the first day of the prediction window with zero latency in epidemiological reporting, representing a best-case scenario for predictive skill.

Across all lead times, we compared the best-fitting model with adjusted formulations, such as exclusion of the yearly random effect and unstratified covariate effects, and corresponding baseline random-effects only models. Next, we explored different schemes for handling the yearly random effect (Text S1) at all lead times. We then systematically altered the architecture of the best-fitting model to evaluate various permutations, exclusively at the 3-month lead time, by iteratively removing or adding fixed effects. Lastly, we tested the inclusion of mobility covariates at all lead times.

Forecast skill was assessed using a range of predictive skill metrics. The Continuous Ranked Probability Score (CRPS) measures the accuracy of probabilistic forecasts for dengue case counts by comparing the predicted cumulative distribution to the observation. The Brier Score (BS) evaluates probabilistic accuracy by calculating the mean squared difference between the outbreak probability and whether an outbreak occurred. The Receiver Operating Characteristic (ROC) curve plots the True Positive Rate (TPR; *sensitivity*), the proportion of outbreak months correctly alerted, against the False Positive Rate (FPR; *1-specificity*), the proportion of non-outbreak months falsely alerted, across all outbreak probability thresholds. The Area Under the ROC Curve (AUC) discriminates between outbreak months and non-outbreak months, with 1.0 representing perfect discrimination. We reported the TPR and FPR at the trigger threshold closest to perfect discrimination. The trigger threshold was calculated nationally for aggregate analysis across all district-months, and subregionally for spatial analysis. An outbreak was defined as observed cases exceeding the historical district-month-specific mean plus two standard deviations of dengue cases, previously identified as the optimal outbreak threshold to predict province-level dengue risk in Vietnam (Colón-González et al., 2021). We additionally ran our analysis with the 75th and 95th percentiles to internally validate our results. All metrics were quantified as aggregates across all district-months and per subregion, with the CRPS and BS additionally quantified per district.

## Data availability

Dengue case count data per district-month may be formally requested by completing an official data access form and obtaining written approval from the General Department of Preventive Medicine, Ministry of Health, Vietnam via the National Institute of Hygiene and Epidemiology). All other data are available from open sources. Temperature and precipitation data were obtained from ERA5-Land (https://cds.climate.copernicus.eu/datasets/reanalysis-era5-land), oceanic indices from NOAA (https://psl.noaa.gov/data), Köppen-Geiger climate classification from Beck et al. (2023) via GloH2O (https://www.gloh2o.org/koppen/), elevation data from Copernicus GLO-90 DEM (https://portal.opentopography.org/raster?opentopoID=OTSDEM.032021.4326.1), census-data data from the Vietnam General Statistics Office (https://www.gso.gov.vn/en/homepage/), land cover data from ESA-CCI (https://www.esa-landcover-cci.org/), and mobility data from Meta AI for Good (https://ai.meta.com/ai-for-good/datasets/movement-distribution-maps/.

## Acknowledgements

This research was supported by the European Union’s Horizon Europe research and innovation programme, E4Warning (101086640). S.B. was supported by Schmidt Science Fellows, in partnership with the Rhodes Trust. R.G. was supported by the Royal Society via a University Research Fellowship (URF\R1\251820). D.L. was supported by the Barcelona Supercomputing Center AI4Science Fellowship programme funded by the Recovery and Resilience Mechanism-Next Generation as part of the Spanish Ministry’s Recovery, Transformation and Resilience Plan. Q.H. and G.T. were supported by the Medical Research Council (MRC) grant ‘Integrating and scaling seasonal climate-driven dengue forecasting’ (MR/Y004663/1).

## References

Alcayna, T., Kellerhaus, F., Tremblay, L., Fletcher, C., Goodermote, R., Santos-Vega, M., Chaves-Gonzalez, J., Bailey, M., Rao, V. B., & Lowe, R. (2025). Integrating anticipatory action in disease outbreak preparedness and response in the humanitarian sector. BMJ Global Health. 10.1136/bmjgh-2024-017721

Banu, S., Guo, Y., Hu, W., Dale, P., Mackenzie, J. S., Mengersen, K., & Tong, S. (2015). Impacts of El Niño Southern Oscillation and Indian Ocean Dipole on dengue incidence in Bangladesh. Scientific Reports, 5(1), 16105. 10.1038/srep16105

Baston, D., ISciences, & LLC. (2025). exactextractr: Fast Extraction from Raster Datasets using Polygons (Version 0.10.1) [Computer software]. https://cran.r-project.org/web/packages/exactextractr/index.html

Beck, H. E., McVicar, T. R., Vergopolan, N., Berg, A., Lutsko, N. J., Dufour, A., Zeng, Z., Jiang, X., van Dijk, A. I. J. M., & Miralles, D. G. (2023). High-resolution (1 km) Köppen-Geiger maps for 1901–2099 based on constrained CMIP6 projections. Scientific Data, 10(1), 724. 10.1038/s41597-023-02549-6

Belman, S., Lefrancq, N., Nzenze, S., Downs, S., du Plessis, M., Lo, S. W., McGee, L., Madhi, S. A., von Gottberg, A., Bentley, S. D., & Salje, H. (2024). Geographical migration and fitness dynamics of Streptococcus pneumoniae. Nature, 631(8020), 386–392. 10.1038/s41586-024-07626-3

Brady, O. J., Smith, D. L., Scott, T. W., & Hay, S. I. (2015). Dengue disease outbreak definitions are implicitly variable. Epidemics, 11, 92–102. 10.1016/j.epidem.2015.03.002

Brook, C. E., Rozins, C., Bohl, J. A., Ahyong, V., Chea, S., Fahsbender, L., Huy, R., Lay, S., Leang, R., Li, Y., Lon, C., Man, S., Oum, M., Northrup, G. R., Oliveira, F., Pacheco, A. R., Parker, D. M., Young, K., Boots, M., … Manning, J. E. (2024). Climate, demography, immunology, and virology combine to drive two decades of dengue virus dynamics in Cambodia. Proceedings of the National Academy of Sciences, 121(36), e2318704121. 10.1073/pnas.2318704121

Chen, Y., Xu, Y., Wang, L., Liang, Y., Li, N., Lourenço, J., Yang, Y., Lin, Q., Wang, L., Zhao, H., Cazelles, B., Song, H., Liu, Z., Wang, Z., Brady, O. J., Cauchemez, S., & Tian, H. (2024). Indian Ocean temperature anomalies predict long-term global dengue trends. Science, 384(6696), 639–646. 10.1126/science.adj4427

Chuang, T.-W., Chaves, L. F., & Chen, P.-J. (2017). Effects of local and regional climatic fluctuations on dengue outbreaks in southern Taiwan. 12(6), e0178698. 10.1371/journal.pone.0178698

Colón-González, F. J., Bastos, L. S., Hofmann, B., Hopkin, A., Harpham, Q., Crocker, T., Amato, R., Ferrario, I., Moschini, F., James, S., Malde, S., Ainscoe, E., Nam, V. S., Tan, D. Q., Khoa, N. D., Harrison, M., Tsarouchi, G., Lumbroso, D., Brady, O. J., & Lowe, R. (2021). Probabilistic seasonal dengue forecasting in Vietnam: A modelling study using superensembles. PLOS Medicine, 18(3), e1003542. 10.1371/journal.pmed.1003542

Díaz, A. R., Rollock, L., Boodram, L.-L. G., Mahon, R., Best, S., Trotman, A., Meerbeeck, C. J. V., Fletcher, C., Dunbar, W., Lippi, C. A., Lührsen, D., Sorensen, C., Muñoz, Á. G., Ryan, S. J., Stewart-Ibarra, A. M., & Lowe, R. (2024). A demand-driven climate services for health implementation framework: A case study for climate-sensitive diseases in Caribbean Small Island Developing States. 3(10), e0000282. 10.1371/journal.pclm.0000282

Djaafara, B. A., Elyazar, I. R. F., Silalahi, F. S. M., Surya, A., Handito, A., Thohir, B., Aryani, D., Kamal, M., Ramadona, A. L., Gunawan, D., Hipokrates, Nisa, A. K., Prianto, E., Samad, I., Sugiarto, A., Fornace, K., Clapham, H. E., Faria, N. R., & Mishra, S. (2026). Dengue transmission heterogeneity across Indonesia’s archipelago: Climate-driven spatiotemporal patterns and policy implications. 20(3), e0014135. 10.1371/journal.pntd.0014135

Do, Q. V., Do, H. X., Do, N. C., & Ngo, A. L. (2020). Changes in Precipitation Extremes across Vietnam and Its Relationships with Teleconnection Patterns of the Northern Hemisphere. Water, 12(6), 1646. 10.3390/w12061646

Doeurk, B., Marcombe, S., Maquart, P.-O., & Boyer, S. (2024). Review of dengue vectors in Cambodia: Distribution, bionomics, vector competence, control and insecticide resistance. Parasites & Vectors, 17(1), 424. 10.1186/s13071-024-06481-5

Duong, C. V., Kang, J. H., Nguyen, V. V., & Bae, Y. J. (2022). Invasion Pattern of Aedes aegypti in the Native Range of Ae. Albopictus in Vietnam Revealed by Biogeographic and Population Genetic Analysis. Insects, 13(12), 1079. 10.3390/insects13121079

European Space Agency. (2024). Copernicus Global Digital Elevation Model. OpenTopography. 10.5069/G9028PQB

Finch, E., Chang, C., Kucharski, A., Sim, S., Ng, L.-C., & Lowe, R. (2025). Climate variation and serotype competition drive dengue outbreak dynamics in Singapore. Nature Communications, 16(1), 11364. 10.1038/s41467-025-66411-6

Fletcher, C., Moirano, G., Alcayna, T., Rollock, L., Meerbeeck, C. J. V., Mahon, R., Trotman, A., Boodram, L.-L., Browne, T., Best, S., Lührsen, D., Diaz, A. R., Dunbar, W., Lippi, C. A., Ryan, S. J., Colón-González, F. J., Stewart-Ibarra, A. M., & Lowe, R. (2025). Compound and cascading effects of climatic extremes on dengue outbreak risk in the Caribbean: An impact-based modelling framework with long-lag and short-lag interactions. The Lancet Planetary Health, 9(8). 10.1016/j.lanplh.2025.06.003

GADM. (2026). *GADM*. https://gadm.org/download_country.html

García-Carreras, B., Yang, B., Grabowski, M. K., Sheppard, L. W., Huang, A. T., Salje, H., Clapham, H. E., Iamsirithaworn, S., Doung-Ngern, P., Lessler, J., & Cummings, D. A. T. (2022). Periodic synchronisation of dengue epidemics in Thailand over the last 5 decades driven by temperature and immunity. PLoS Biology, 20(3), e3001160. 10.1371/journal.pbio.3001160

Gibb, R., Colón-González, F. J., Lan, P. T., Huong, P. T., Nam, V. S., Duoc, V. T., Hung, D. T., Dong, N. T., Chien, V. C., Trang, L. T. T., Kien Quoc, D., Hoa, T. M., Tai, N. H., Hang, T. T., Tsarouchi, G., Ainscoe, E., Harpham, Q., Hofmann, B., Lumbroso, D., … Lowe, R. (2023). Interactions between climate change, urban infrastructure and mobility are driving dengue emergence in Vietnam. Nature Communications, 14(1), 8179. 10.1038/s41467-023-43954-0

Gloria-Soria, A., Armstrong, P. M., Powell, J. R., & Turner, P. E. (2017). Infection rate of Aedes aegypti mosquitoes with dengue virus depends on the interaction between temperature and mosquito genotype. Proceedings of the Royal Society B: Biological Sciences, 284(1864), 20171506. 10.1098/rspb.2017.1506

Gómez-Vargas, W., Ríos-Tapias, P. A., Marin-Velásquez, K., Giraldo-Gallo, E., Segura-Cardona, A., & Arboleda, M. (2024). Density of Aedes aegypti and dengue virus transmission risk in two municipalities of Northwestern Antioquia, Colombia. 19(1), e0295317. 10.1371/journal.pone.0295317

Harish, V., Colón-González, F. J., Moreira, F. R. R., Gibb, R., Kraemer, M. U. G., Davis, M., Reiner, R. C., Pigott, D. M., Perkins, T. A., Weiss, D. J., Bogoch, I. I., Vazquez-Prokopec, G., Saide, P. M., Barbosa, G. L., Sabino, E. C., Khan, K., Faria, N. R., Hay, S. I., Correa-Morales, F., … Brady, O. J. (2024). Human movement and environmental barriers shape the emergence of dengue. Nature Communications, 15(1), 4205. 10.1038/s41467-024-48465-0

Hien, N. T., Anh, D. D., Le, N. H., Yen, N. T., Phong, T. V., Nam, V. S., Duong, T. N., Nguyen, N. B., Huong, D. T. T., Hung, L. Q., Trinh, C. N. T., Hoang, N. V., Mai, V. Q., Nghia, L. T., Dong, N. T., Tho, L. H., Kutcher, S., Hurst, T. P., Montgomery, J. L., … Ryan, P. A. (2022). Environmental factors influence the local establishment of Wolbachia in Aedes aegypti mosquitoes in two small communities in central Vietnam. Gates Open Research, 5, 147. 10.12688/gatesopenres.13347.2

Houatthongkham, S., Kim, J. H., Khamphaphongphane, B., Xangsayarath, P., Kim, J.-H., & Kim, S. H. (2026). Climate drivers and winter constraints of dengue epidemics: A 10-year epidemiological perspective study in the Lao People’s Democratic Republic. Infectious Diseases of Poverty, 15(1), 46. 10.1186/s40249-026-01438-5

Joanne, S., Vythilingam, I., Teoh, B.-T., Leong, C.-S., Tan, K.-K., Wong, M.-L., Yugavathy, N., & AbuBakar, S. (2017). Vector competence of Malaysian Aedes albopictus with and without Wolbachia to four dengue virus serotypes. Tropical Medicine & International Health, 22(9), 1154–1165. 10.1111/tmi.12918

Kiang, M. V., Santillana, M., Chen, J. T., Onnela, J.-P., Krieger, N., Engø-Monsen, K., Ekapirat, N., Areechokchai, D., Prempree, P., Maude, R. J., & Buckee, C. O. (2021). Incorporating human mobility data improves forecasts of Dengue fever in Thailand. Scientific Reports, 11(1), 923. 10.1038/s41598-020-79438-0

Kim Lien, P. T., Duoc, V. T., Gavotte, L., Cornillot, E., Nga, P. T., Briant, L., Frutos, R., & Duong, T. N. (2015). Role of *Aedes aegypti* and *Aedes albopictus* during the 2011 dengue fever epidemics in Hanoi, Vietnam. Asian Pacific Journal of Tropical Medicine, 8(7), 543–548. 10.1016/j.apjtm.2015.06.009

Kobayashi, D., Kai, I., Faizah, A. N., Moi, M. L., Tajima, S., Takasaki, T., Sasaki, T., & Isawa, H. (2023). Comparative analysis of the susceptibility of Aedes aegypti and Japanese Aedes albopictus to all dengue virus serotypes. Tropical Medicine and Health, 51(1), 61. 10.1186/s41182-023-00553-5

Kramer, I. M., Vereecken, S., Vanslembrouck, A., Smekens, Y., de Witte, J., Vielma, S., Niamir, A., & Müller, R. (2026). Heatwaves Constrain the Future Persistence of Mosquito Vectors in Europe. Global Change Biology, 32, e70876. 10.1111/gcb.70876

Le, A. T., Pham, H. T., Vu, K.-D., Khuy, C. M., Lien, T. H. M., Duyen, P. T. T., Le, H. H. T. C., Hung, T. M., Phung, D., & Nam, V. S. (2026). Seroprevalence of dengue virus infection in people residing in Vietnam: A systematic review and meta-analysis. Public Health, 256, 106300. 10.1016/j.puhe.2026.106300

Lee, S. A., Economou, T., Catão, R. de C., Barcellos, C., & Lowe, R. (2021). The impact of climate suitability, urbanisation, and connectivity on the expansion of dengue in 21st century Brazil. 15(12), e0009773. 10.1371/journal.pntd.0009773

Liu, X., Huang, Y., Xu, X., Li, X., Li, X., Ciais, P., Lin, P., Gong, K., Ziegler, A. D., Chen, A., Gong, P., Chen, J., Hu, G., Chen, Y., Wang, S., Wu, Q., Huang, K., Estes, L., & Zeng, Z. (2020). High-spatiotemporal-resolution mapping of global urban change from 1985 to 2015. Nature Sustainability, 3(7), 564–570. 10.1038/s41893-020-0521-x

Lowe, R., Gasparrini, A., Meerbeeck, C. J. V., Lippi, C. A., Mahon, R., Trotman, A. R., Rollock, L., Hinds, A. Q. J., Ryan, S. J., & Stewart-Ibarra, A. M. (2018). Nonlinear and delayed impacts of climate on dengue risk in Barbados: A modelling study. 15(7), e1002613. 10.1371/journal.pmed.1002613

Lowe, R., Lee, S. A., O’Reilly, K. M., Brady, O. J., Bastos, L., Carrasco-Escobar, G., de Castro Catão, R., Colón-González, F. J., Barcellos, C., Carvalho, M. S., Blangiardo, M., Rue, H., & Gasparrini, A. (2021). Combined effects of hydrometeorological hazards and urbanisation on dengue risk in Brazil: A spatiotemporal modelling study. The Lancet Planetary Health, 5(4), e209–e219. 10.1016/S2542-5196(20)30292-8

Meta. (2024). Movement Distribution Maps | AFG Dataset | AI at Meta. https://ai.meta.com/ai-for-good/datasets/movement-distribution-maps/

Milà, C., Moirano, G., Kawiecki, A. B., & Lowe, R. (2025). GHRmodel: Bayesian Hierarchical Modelling of Spatio-Temporal Health Data (Version 0.1.1) [Computer software]. https://cran.r-project.org/web/packages/GHRmodel/index.html

Mordecai, E. A., Caldwell, J. M., Grossman, M. K., Lippi, C. A., Johnson, L. R., Neira, M., Rohr, J. R., Ryan, S. J., Savage, V., Shocket, M. S., Sippy, R., Stewart Ibarra, A. M., Thomas, M. B., & Villena, O. (2019). Thermal biology of mosquito-borne disease. Ecology Letters, 22(10), 1690–1708. 10.1111/ele.13335

National Statistics Office of Vietnam. (2010). The 2009 Vietnam Population and Housing census: Completed results. https://www.nso.gov.vn/en/data-and-statistics/2019/03/the-2009-vietnam-population-and-housing-census-completed-results/

National Statistics Office of Vietnam. (2020). Completed results of the 2019 Viet Nam population and housing census. https://www.nso.gov.vn/en/data-and-statistics/2020/11/completed-results-of-the-2019-viet-nam-population-and-housing-census/

Nguyen, C. T., Moi, M. L., Le, T. Q. M., Nguyen, T. T. T., Vu, T. B. H., Nguyen, H. T., Pham, T. T. H., Le, T. H. T., Nguyen, L. M. H., Phu Ly, M. H., Ng, C. F. S., Takemura, T., Morita, K., & Hasebe, F. (2020). Prevalence of Zika virus neutralizing antibodies in healthy adults in Vietnam during and after the Zika virus epidemic season: A longitudinal population-based survey. BMC Infectious Diseases, 20(1), 332. 10.1186/s12879-020-05042-2

Nguyen, L. T., Le, H. X., Nguyen, D. T., Ho, H. Q., & Chuang, T.-W. (2020). Impact of Climate Variability and Abundance of Mosquitoes on Dengue Transmission in Central Vietnam. International Journal of Environmental Research and Public Health, 17(7), 2453. 10.3390/ijerph17072453

Nguyen, T. H., Nguyen, H. L., Nguyen, T. Y., Vu, S. N., Tran, N. D., Le, T. N., Vien, Q. M., Bui, T. C., Le, H. T., Kutcher, S., Hurst, T. P., Duong, T. T. H., Jeffery, J. A. L., Darbro, J. M., Kay, B. H., Iturbe-Ormaetxe, I., Popovici, J., Montgomery, B. L., Turley, A. P., … Hoffmann, A. A. (2015). Field evaluation of the establishment potential of wmelpop Wolbachia in Australia and Vietnam for dengue control. Parasites & Vectors, 8(1), 563. 10.1186/s13071-015-1174-x

Nguyen-Le, D., Ngo-Duc, T., & Matsumoto, J. (2024). The teleconnection of the two types of ENSO and Indian Ocean Dipole on Southeast Asian autumn rainfall anomalies. Climate Dynamics, 62(6), 1–23. 10.1007/s00382-024-07163-9

Phan-Van, T., Nguyen-Ngoc-Bich, P., Ngo-Duc, T., Vu-Minh, T., Le, P. V. V., Trinh-Tuan, L., Nguyen-Thi, T., Pham-Thanh, H., & Tran-Quang, D. (2022). Drought over Southeast Asia and Its Association with Large-Scale Drivers. 35(15), 4959–4978. 10.1175/JCLI-D-21-0770.1

Quan, T. M., Phuong, H. T., Vy, N. H. T., Thanh, N. T. L., Lien, N. T. N., Hong, T. T. K., Dung, P. N., Chau, N. V. V., Boni, M. F., & Clapham, H. E. (2018). Evidence of previous but not current transmission of chikungunya virus in southern and central Vietnam: Results from a systematic review and a seroprevalence study in four locations. 12(2), e0006246. 10.1371/journal.pntd.0006246

Rue, H., Martino, S., & Chopin, N. (2009). Approximate Bayesian Inference for Latent Gaussian models by using Integrated Nested Laplace Approximations. Journal of the Royal Statistical Society Series B: Statistical Methodology, 71(2), 319–392. 10.1111/j.1467-9868.2008.00700.x

Sáez-Llorens, X., DeAntonio, R., Low, J. G. H., Kosalaraksa, P., Dean, H., Sharma, M., Tricou, V., & Biswal, S. (2025). TAK-003: Development of a tetravalent dengue vaccine. Expert Review of Vaccines, 24(1), 324–338. 10.1080/14760584.2025.2490295

Salje, H., Wesolowski, A., Brown, T. S., Kiang, M. V., Berry, I. M., Lefrancq, N., Fernandez, S., Jarman, R. G., Ruchusatsawat, K., Iamsirithaworn, S., Vandepitte, W. P., Suntarattiwong, P., Read, J. M., Klungthong, C., Thaisomboonsuk, B., Engø-Monsen, K., Buckee, C., Cauchemez, S., & Cummings, D. A. T. (2021). Reconstructing unseen transmission events to infer dengue dynamics from viral sequences. Nature Communications, 12(1), 1810. 10.1038/s41467-021-21888-9

Schmidt, W.-P., Suzuki, M., Thiem, V. D., White, R. G., Tsuzuki, A., Yoshida, L.-M., Yanai, H., Haque, U., Tho, L. H., Anh, D. D., & Ariyoshi, K. (2011). Population Density, Water Supply, and the Risk of Dengue Fever in Vietnam: Cohort Study and Spatial Analysis. 8(8), e1001082. 10.1371/journal.pmed.1001082

Stewart-Ibarra, A. M., Ryan, S. J., Beltrán, E., Mejía, R., Silva, M., & Muñoz, Á. (2013). Dengue Vector Dynamics (Aedes aegypti) Influenced by Climate and Social Factors in Ecuador: Implications for Targeted Control. 8(11), e78263. 10.1371/journal.pone.0078263

Tran, H. P., Huynh, T. T. T., Nguyen, Y. T., Kutcher, S., O’Rourke, P., Marquart, L., Ryan, P. A., & Kay, B. H. (2012). Low Entomological Impact of New Water Supply Infrastructure in Southern Vietnam, with Reference to Dengue Vectors. The American Journal of Tropical Medicine and Hygiene, 87(4), 631–639. 10.4269/ajtmh.2012.12-0335

VietNamNet Global. (2025). Vietnam to consolidate into 34 provinces and 3,321 communes by 2025. https://vietnamnet.vn/en/vietnam-to-consolidate-into-34-provinces-and-3-321-communes-by-2025-2399871.html

Vu, H. H., Okumura, J., Hashizume, M., Tran, D. N., & Yamamoto, T. (2014). Regional Differences in the Growing Incidence of Dengue Fever in Vietnam Explained by Weather Variability. Tropical Medicine and Health, 42(1), 25–33. 10.2149/tmh.2013-24

Wang, D., Wu, X., Chen, J., & Zhao, L. (2026). Urbanization, infrastructure, and mosquito thermal suitability shape the causal effects of extreme weather on dengue transmission. 10.59717/j.xinn-med.2026.100188

Wang, Y., Chong, K. C., & Ren, C. (2024). Impact of compound warm and wet events on dengue fever infection in South and Southeast Asian countries. Environmental Research, 263, 120091. 10.1016/j.envres.2024.120091

Wanthanaporn, U., Supit, I., Hove, B. van, & Hutjes, R. W. A. (2025). Analysis of Seasonal Climate and Streamflow Forecast Performance for Mainland Southeast Asia. 26(12), 1849–1866. 10.1175/JHM-D-25-0092.1

Wartel, T. A., Prayitno, A., Hadinegoro, S. R. S., Capeding, M. R., Thisyakorn, U., Tran, N. H., Moureau, A., Bouckenooghe, A., Nealon, J., & Taurel, A. F. (2016). Three Decades of Dengue Surveillance in Five Highly Endemic South East Asian Countries: A Descriptive Review. 29(1), 7–16. 10.1177/1010539516675701

Weisheimer, A., & Palmer, T. N. (2014). On the reliability of seasonal climate forecasts. Journal of the Royal Society Interface, 11(96), 20131162. 10.1098/rsif.2013.1162

Whitehorn, J., Kien, D. T. H., Nguyen, N. M., Nguyen, H. L., Kyrylos, P. P., Carrington, L. B., Tran, C. N. B., Quyen, N. T. H., Thi, L. V., Le Thi, D., Truong, N. T., Luong, T. T. H., Nguyen, C. V. V., Wills, B., Wolbers, M., & Simmons, C. P. (2015). Comparative Susceptibility of *Aedes albopictus* and *Aedes aegypti* to Dengue Virus Infection After Feeding on Blood of Viremic Humans: Implications for Public Health. Journal of Infectious Diseases, 212(8), 1182–1190. 10.1093/infdis/jiv173

World Health Organization. (2024). Dengue and severe dengue. https://www.who.int/news-room/fact-sheets/detail/dengue-and-severe-dengue

Worldometer. (2026a). Largest Countries in the World by Area. https://www.worldometers.info/geography/largest-countries-in-the-world/

Worldometer. (2026b). Population by Country (2026). https://www.worldometers.info/world-population/population-by-country/

Zscheischler, J., Martius, O., Westra, S., Bevacqua, E., Raymond, C., Horton, R. M., van den Hurk, B., AghaKouchak, A., Jézéquel, A., Mahecha, M. D., Maraun, D., Ramos, A. M., Ridder, N. N., Thiery, W., & Vignotto, E. (2020). A typology of compound weather and climate events. Nature Reviews Earth & Environment, 1(7), 333–347. 10.1038/s43017-020-0060-z

Zscheischler, J., Westra, S., van den Hurk, B. J. J. M., Seneviratne, S. I., Ward, P. J., Pitman, A., AghaKouchak, A., Bresch, D. N., Leonard, M., Wahl, T., & Zhang, X. (2018). Future climate risk from compound events. Nature Climate Change, 8(6), 469–477. 10.1038/s41558-018-0156-3

