## Supplementary Appendix for "Compound climate extremes, socioeconomic conditions and human mobility influence dengue dynamics heterogeneously across Vietnam"

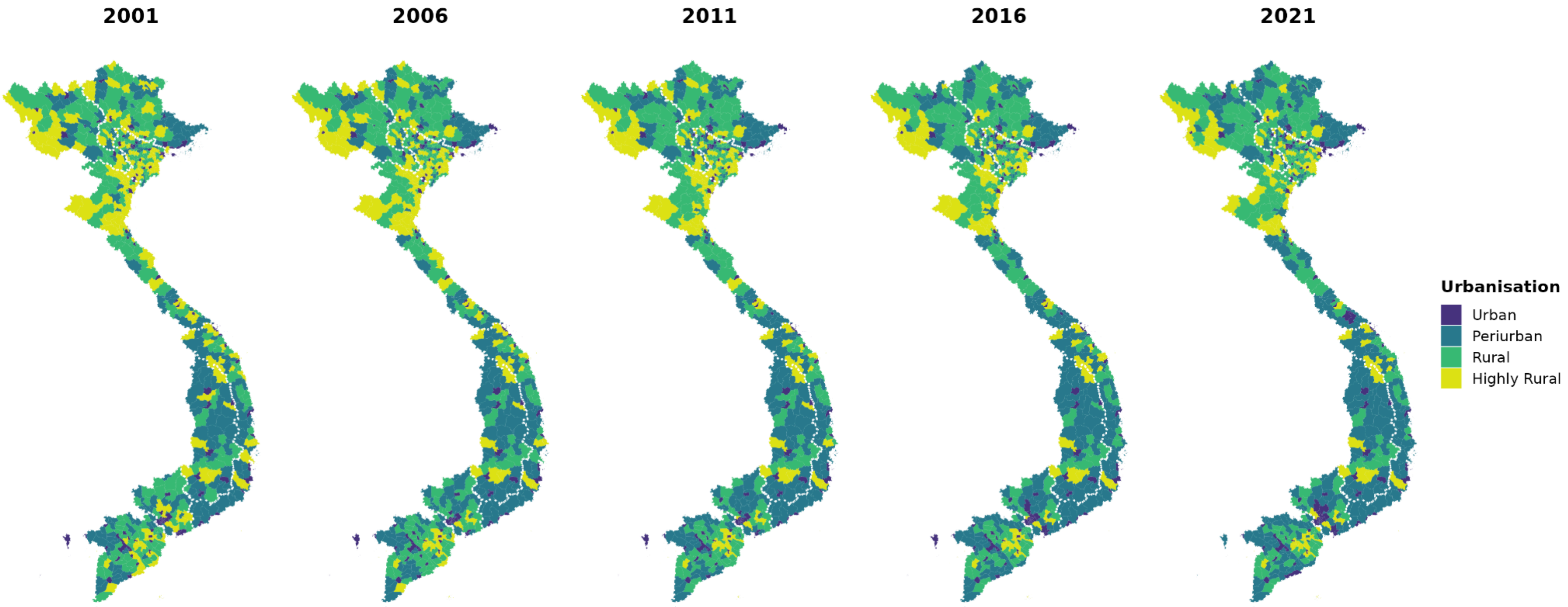


**Figure S1. District-level urbanisation level at five-year intervals from 2001 to 2021.** Each district was assigned an annual urbanisation level using quartiles of the proportion of inhabitants living in urban areas across the study period, defined as Highly Rural [0.0, 5.0]%, Rural [5.1, 10.5]%, Periurban [10.6, 52.9]%, and Urban [53.0, 100.0]%. Urban populations were derived from the Vietnam Population and Housing Censuses, interpolated to annual estimates. Over the 20-year period, 503 districts retained the same classification, whereas 129 changed once, 27 changed twice, and 11 changed three times. Maps only show the 670 districts modelled.

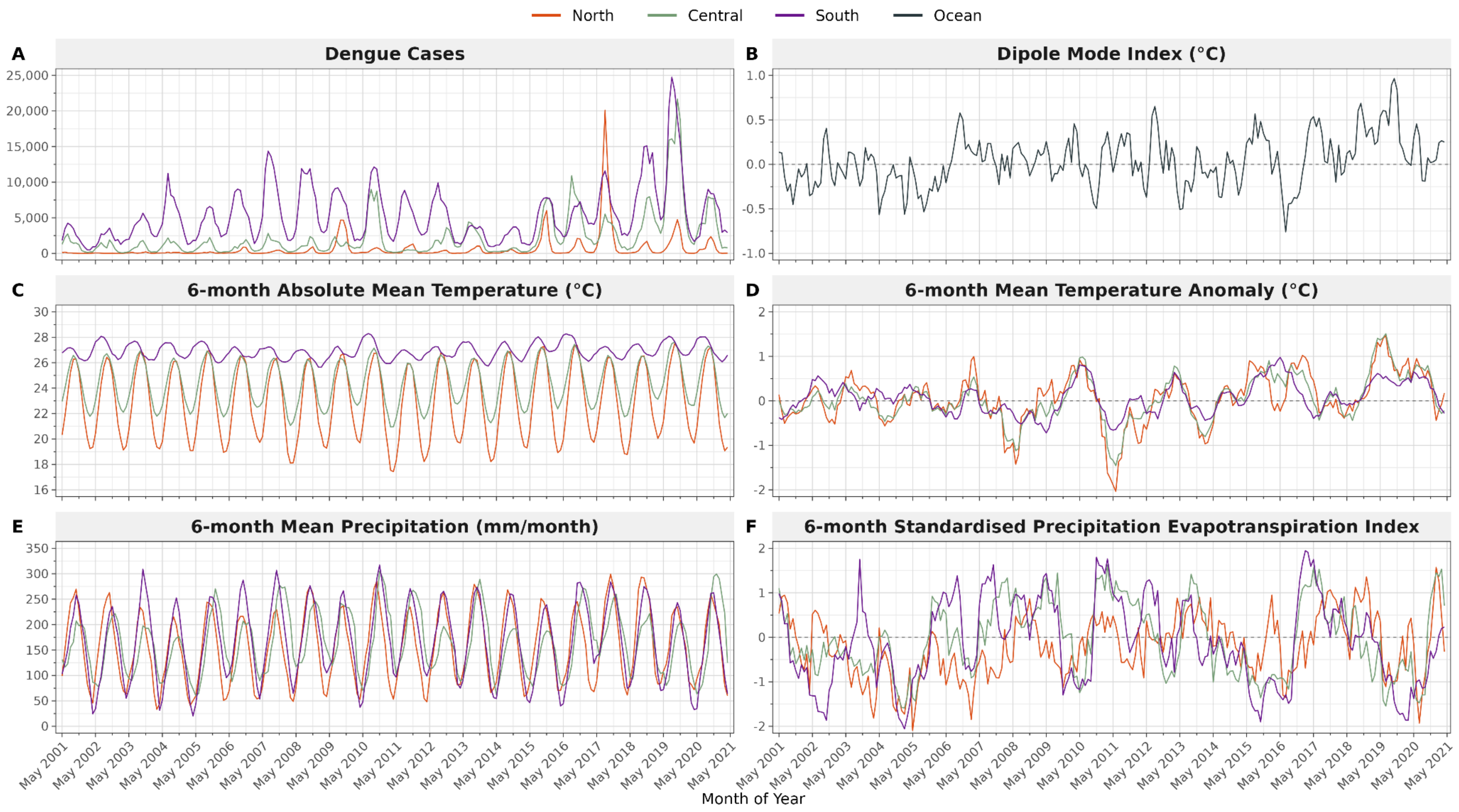


**Figure S2. Regional monthly dengue cases and 6-month averaged climate exposures, and the Dipole Mode Index, from May 2001 to April 2021.** (A) Total monthly dengue cases per region (North, Central and South), (B) Dipole Mode Index which represents the Indian Ocean sea surface temperature gradient (°C), (C) 6-month absolute mean temperature per region (°C), (D) 6-month mean temperature anomaly per region (°C), (E) 6-month mean precipitation (mm/month), and (F) 6-month standardised precipitation evapotranspiration index (SPEI-6). (C)-(F) are population-weighted means of district-level values, calculated per month.

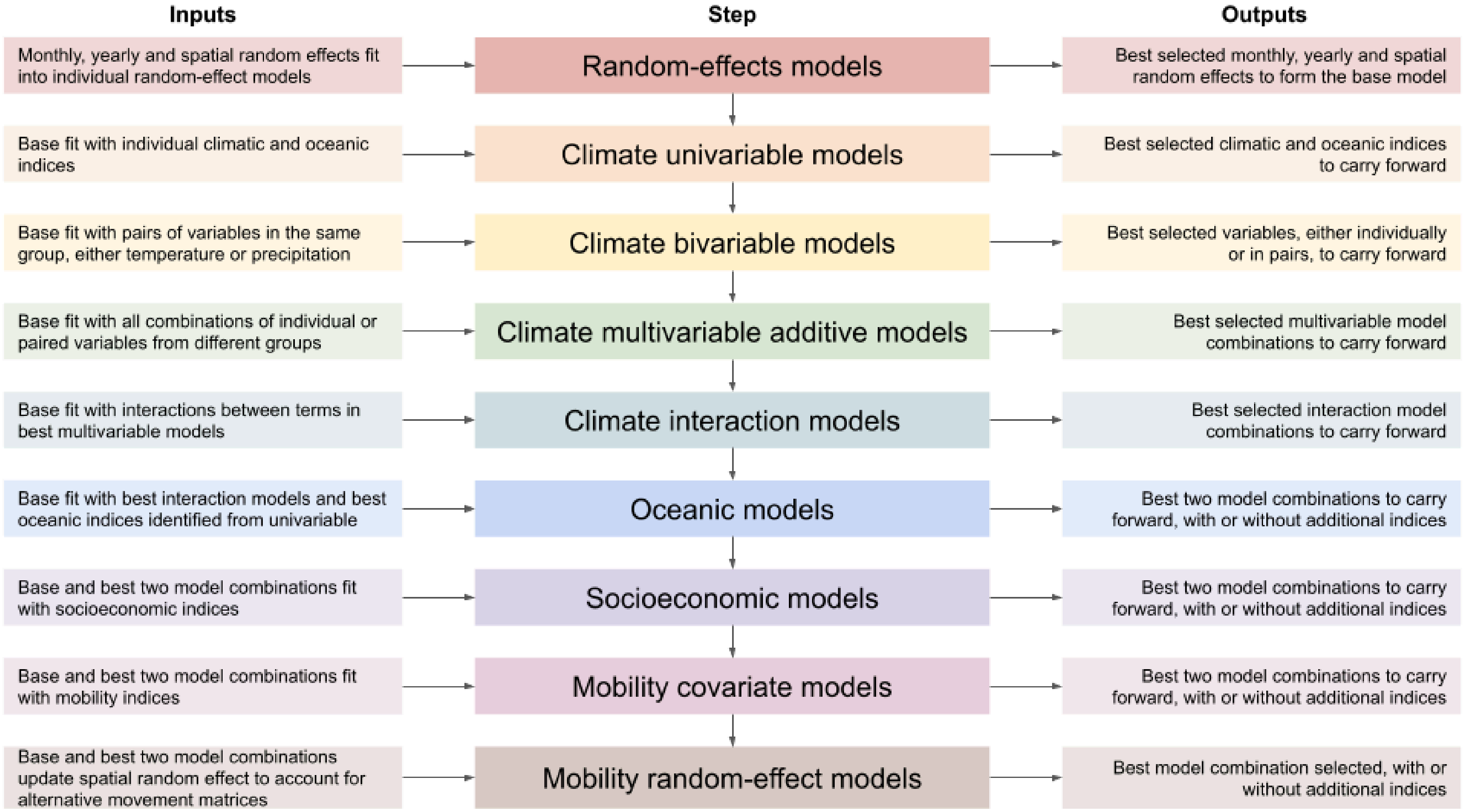


**Figure S3. Stepwise model development framework.** Sequence of model selection stages (centre), with the inputs carried into each stage (left) and the outputs carried forward from each stage (right). At each stage, candidates were compared using goodness-of-fit metrics alongside credible effect sizes and reduced random-effect variance relative to the baseline (base) model.

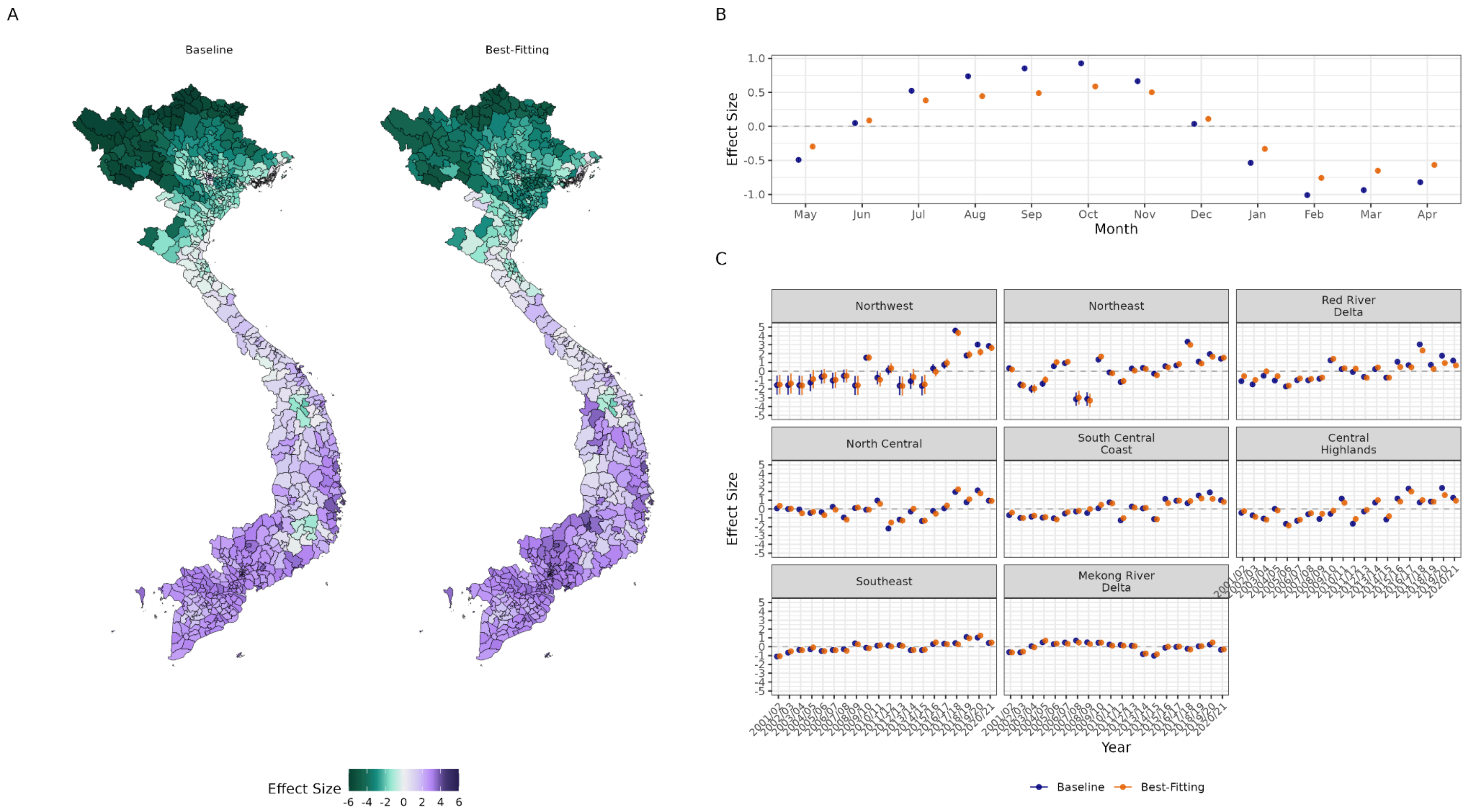


**Figure S4. Spatial, monthly and yearly random effects for the baseline and best-fitting model.** (A) District-level spatial random effects, including structured and unstructured components. (B) Monthly random effects from May to April. (C) Yearly random effects replicated by subregion from 2001/02 to 2020/21, where dengue seasons span May to April. All effects are shown as posterior medians on the log scale. Vertical lines in (B) and (C) represent the 95% credible intervals for the baseline random-effects only model (blue) and the best-fitting socio-climatic model (orange).

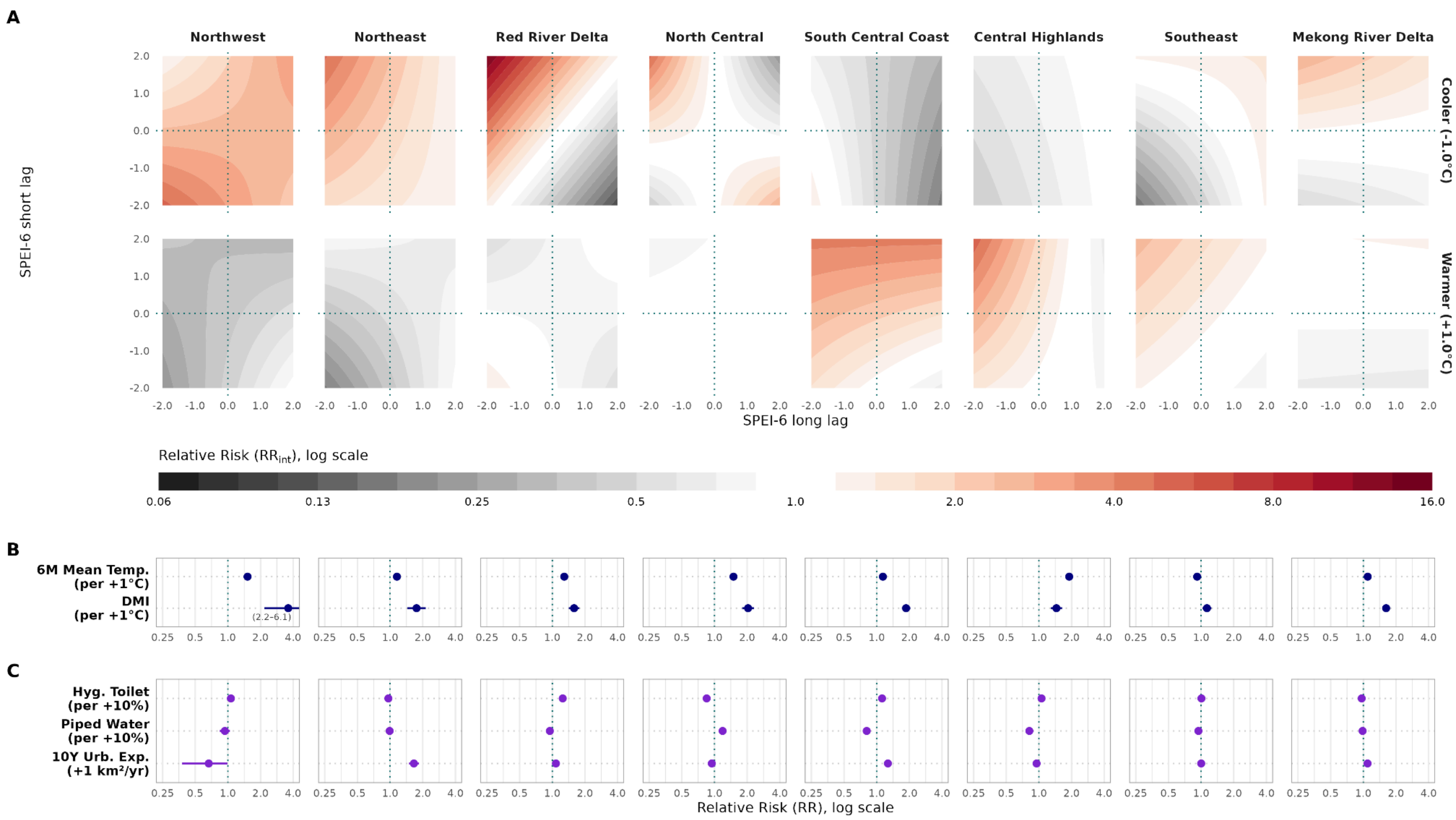


**Figure S5. Subregionally stratified effects of compound climate extremes and additive climatic, oceanic and socioeconomic factors on dengue relative risk with a three-way interaction.** Effects are estimated for the best three-way interaction model with covariates stratified by eight subregions (columns). (A) Relative risk of dengue ($RR_{int}$) across the three-way interaction between the 6-month mean temperature anomaly (lag 0 months), and the SPEI-6 long-lag (lag 4 months) and short-lag (lag 0 months). Surfaces show the posterior mean $RR_{int}$ over approximately ±2 standard deviations of the long-lag SPEI-6 (x-axis), short-lag SPEI-6 (y-axis), and temperature anomaly (rows) for -1.0°C cool (top) and +1.0°C warm (bottom) relative to zero conditions. (B) Relative risk (RR, mean + 95% CrI) of the 6-month absolute mean temperature (lag 0 months), and the DMI (lag 4 months). (C) Relative risk (RR, mean + 95% CrI) of hygienic toilet access, piped water access, and the 10-year urban expansion rate. The temperature anomaly modified the compound response heterogeneously across subregions, with cooler-than-normal conditions elevating dengue risk in the Northwest, Northeast, Red River Delta, North Central, and Mekong River Delta, and warmer-than-normal conditions in the South Central Coast, Central Highlands, and Southeast.

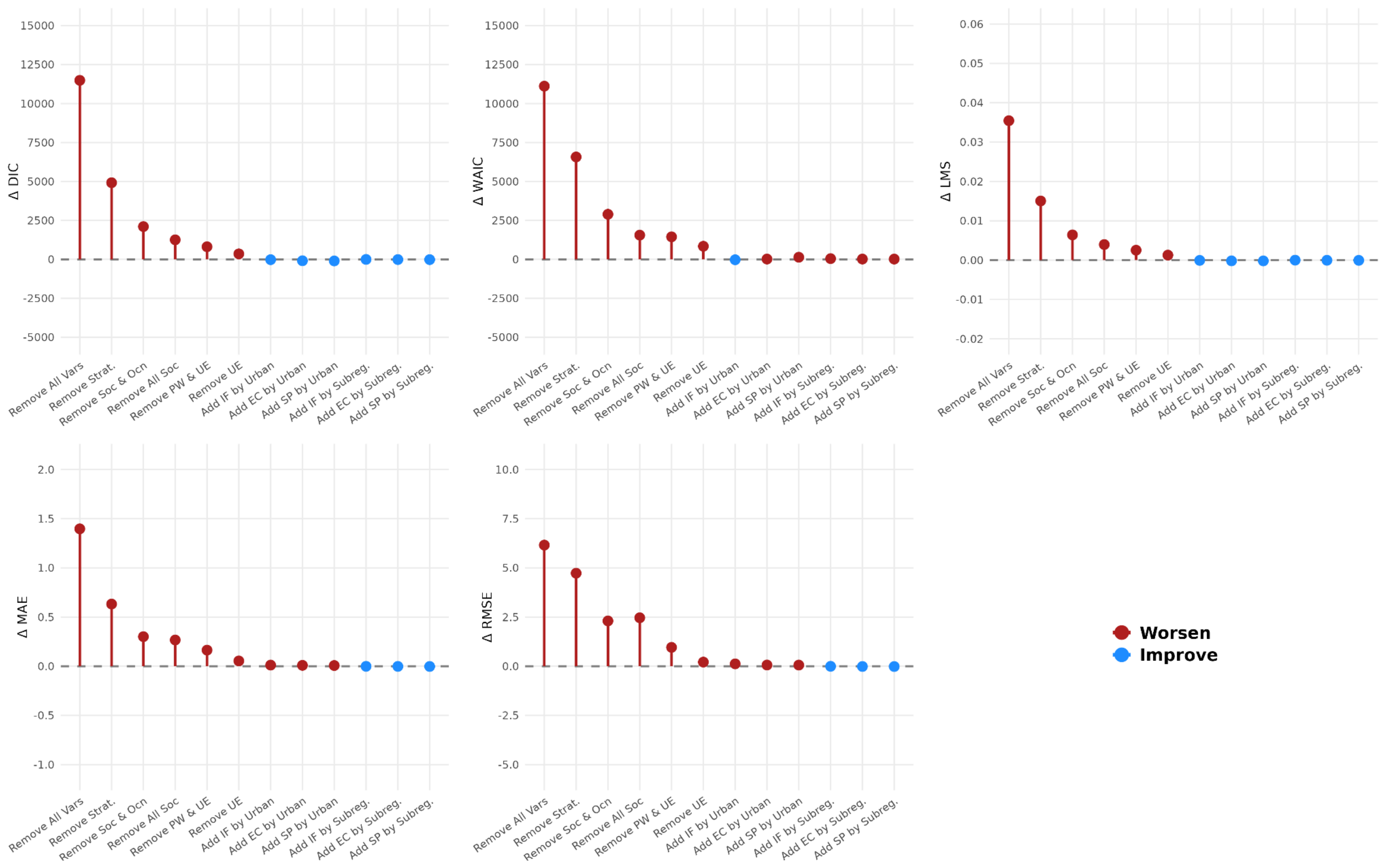


**Figure S6. Change in goodness-of-fit metrics for model variants relative to the best-fitting model.** Difference in the Deviance Information Criterion (DIC), Watanabe-Akaike Information Criterion (WAIC), Log Mean Score (LMS), Mean Absolute Error (MAE), and Root Mean Squared Error (RMSE) between each model variant and the best-fitting model. Variants either remove or add covariates to the best-fitting model. Positive values (red) indicate a worse fit, and negative values (blue) improved fit. Vars = variables; Strat. = spatial stratification (i.e. random slopes); Soc = socioeconomic variables; Ocn = oceanic variables; PW = piped water access; UE = 10-year urban expansion rate; IF = inflow; EC = conditional entropy; SP = staying probability. Mobility covariates (IF, EC, SP) are added as random slopes stratified either by urbanisation level (Urban) or by subregion (Subregion).

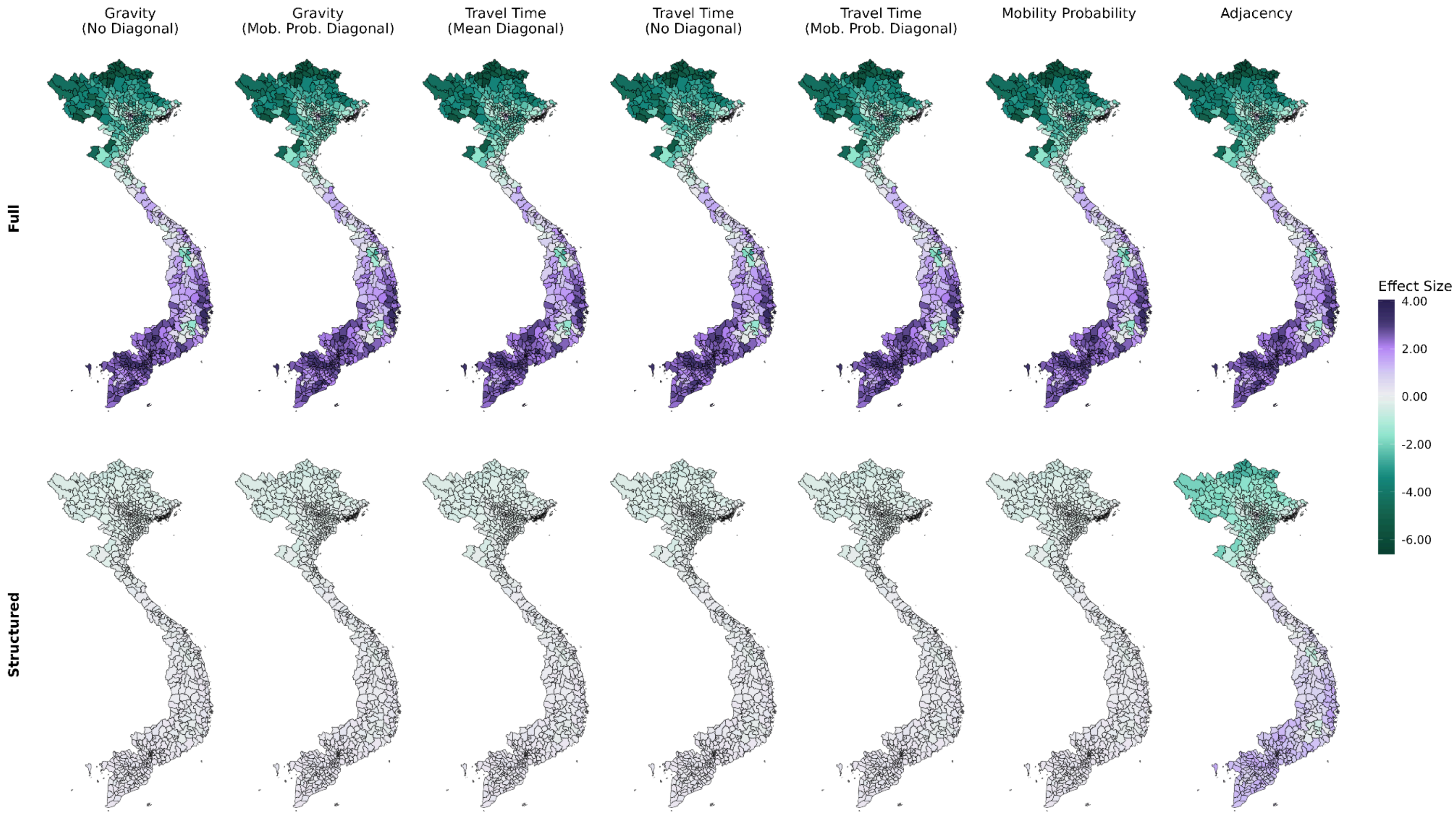


**Figure S7. District-level spatial random effects under alternative neighbourhood structures.** Posterior mean of the BYM2 spatial random effect for seven specifications of the district neighbourhood matrix (from left to right): (i) gravity model with no diagonal, (ii) gravity model with the mobility probability diagonal, (iii) travel time with mean travel times, (iv) travel time with no diagonal, (v) travel time with the mobility probability diagonal, (vi) mobility probability, and (vii) spatial adjacency. Top row (Full) shows the total random effect, combining the structured and unstructured spatial components, and bottom row (Structured) shows the structured component only. Effects are on the log scale, with purple indicating higher and green lower dengue risk. The models are otherwise identical, including an intercept, monthly random effect, and yearly random effect. Maps only show the 670 districts modelled.

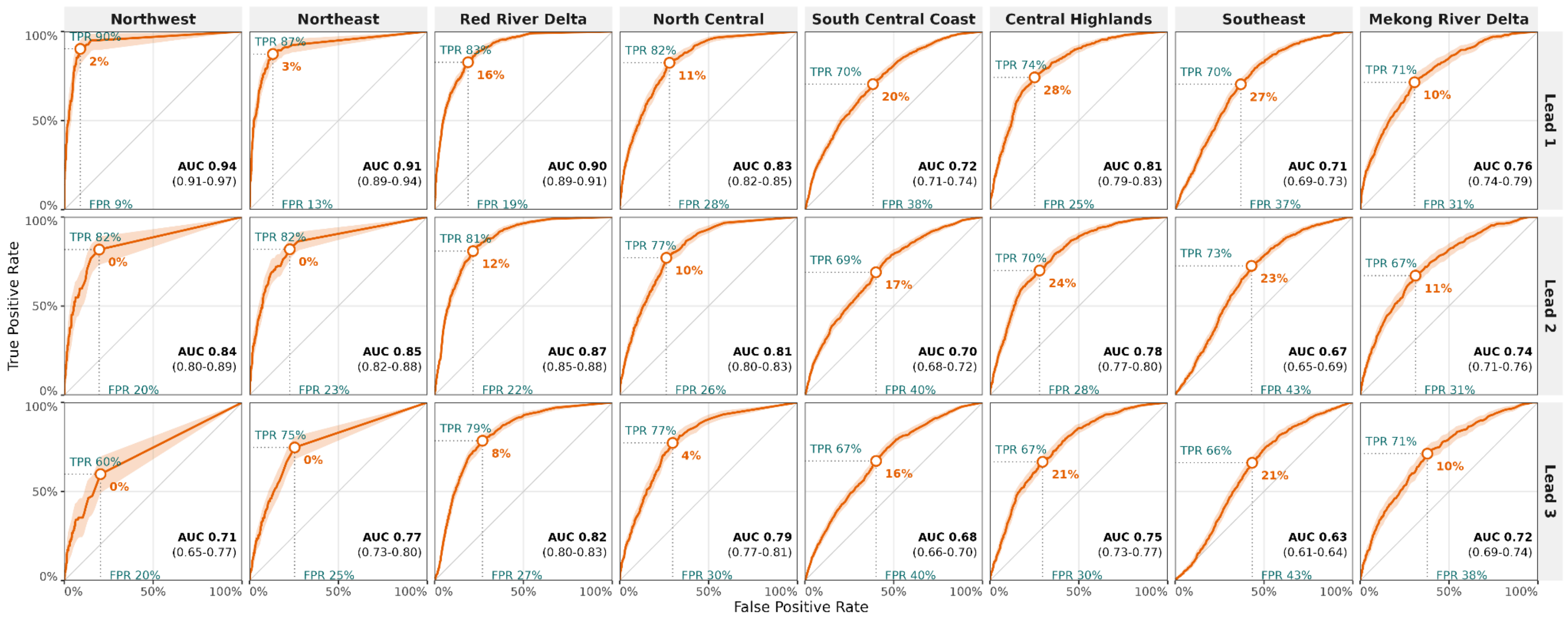


**Figure S8. Subregional discrimination of district-level dengue outbreaks at lead times of 1 to 3 months.** Receiver Operating Characteristic (ROC) curves for model $M_1$ under rolling-origin cross-validation from May 2016 to April 2021 per subregion (columns) per lead time (rows). A district-month is classified as an outbreak when observed cases exceed the historical mean plus two standard deviations. Curves pool all districts and months within a subregion. The solid line represents the mean and shading the 95% confidence interval of the True Positive Rate (TPR) at fixed False Positive Rate (FPR), obtained from 500 bootstrap replicates. The circle marks the trigger probability threshold which minimises the distance to the top-left corner (perfect discrimination), labelled in orange. The TPR and FPR at the trigger threshold are labelled in turquoise. The Area Under the ROC Curve (AUC) is stated with its 95% confidence interval.

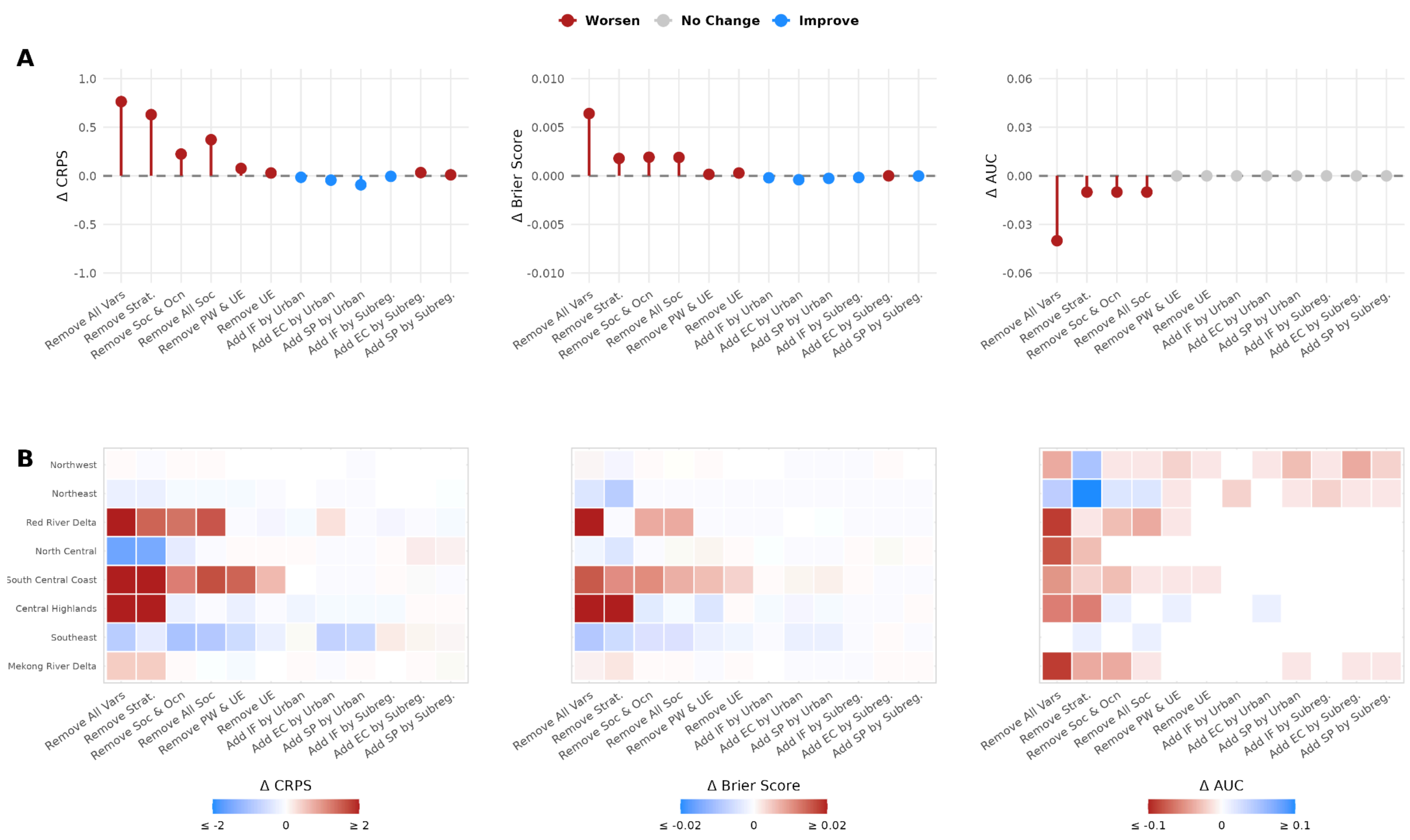


**Figure S9. Change in predictive skill for model variants relative to the best-fitting model at the 3-month lead.** Difference in the Continuous Ranked Probability Score (CRPS), Brier Score (BS), and Area Under the ROC Curve (AUC) between each model variant and the best-fitting model ($M_1$) under rolling-origin cross-validation from May 2016 to April 2021 at a 3-month lead. (A) Differences aggregated nationally. (B) Differences aggregated by subregion. Variants either remove or add covariates to the best-fitting model. Lower CRPS and Brier Score, and higher AUC, indicate improved predictive skill (blue), and the inverse worse predictive skill (red) relative to $M_1$. Vars = variables; Strat. = spatial stratification (i.e. random slopes); Soc = socioeconomic variables; Ocn = oceanic variables; PW = piped water access; UE = 10-year urban expansion rate; IF = inflow; EC = conditional entropy; SP = staying probability. Mobility covariates are added as random slopes stratified either by urbanisation level (Urban) or by subregion (Subregion).

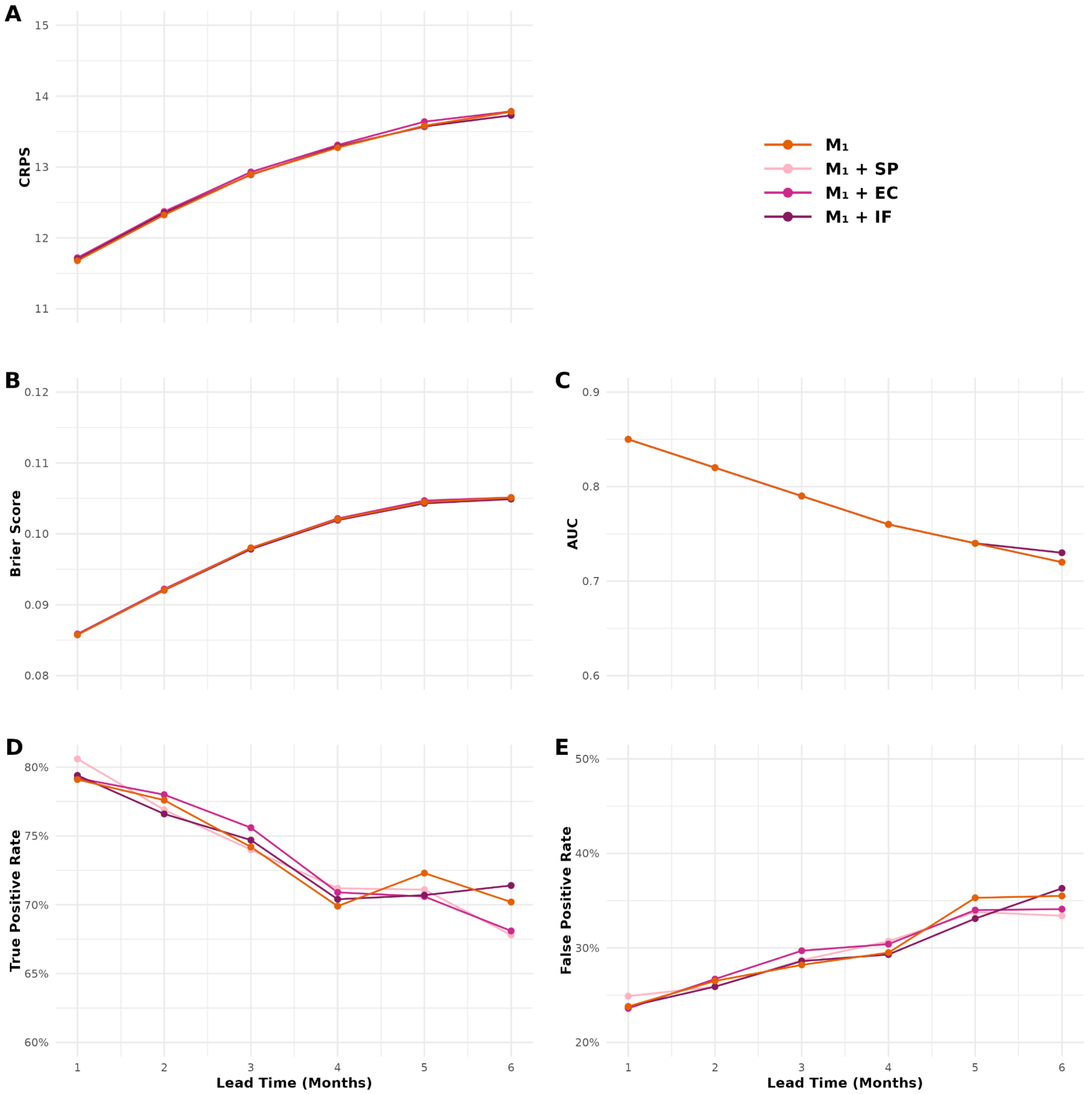


**Figure S10. Predictive skill over all district-months for derivations of the $M_1$ model with mobility covariates at lead times of 1 to 6 months.** Models were evaluated by rolling-origin cross-validation over May 2016 to April 2021. Panels show (A) Continuous Ranked Probability Score (CRPS) for case estimation, (B) Brier Score for probabilistic accuracy, (C) Area Under the Receiver Operating Characteristic Curve (AUC) for outbreak detection, (D) True Positive Rate, and (E) False Positive Rate. Lower values indicate better performance for the CRPS, Brier Score and the False Positive Rate, whereas higher values indicate better performance for the AUC and True Positive Rate. $M_1$ represents the best-fitting model with socio-climatic covariates stratified by subregion and spatial, monthly, and yearly random effects. $M_1$ + SP is the same formulation as $M_1$ adding staying probability stratified by urbanisation level. $M_1$ + EC is the same formulation as $M_1$ adding conditional entropy stratified by urbanisation level. $M_1$ + IF is the same formulation as $M_1$ adding inflow stratified by urbanisation level. This figure shows that mobility covariates have minimal impact on national predictive skill.

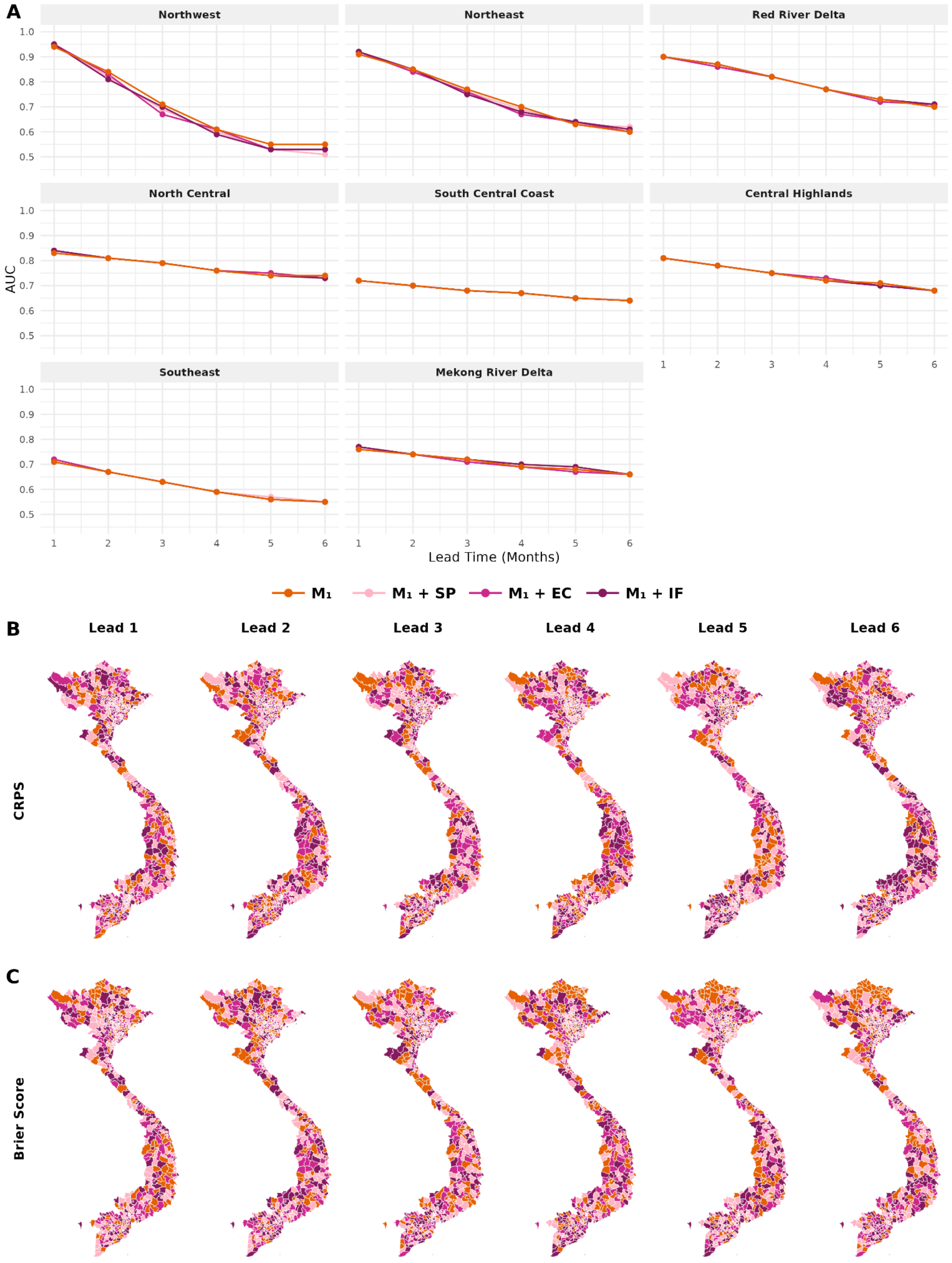


**Figure S11. Subregional and district-level predictive skill for derivations of model $M_1$ with mobility covariates across lead times of 1 to 6 months.** (A) AUC by subregion and lead time for the four models. (B-C) Best-performing model per district by (B) CRPS and (C) Brier Score, at each lead time (columns) coloured by model. $M_1$ represents the best-fitting model with socio-climatic covariates stratified by subregion and spatial, monthly, and yearly random effects. $M_1$ + SP is the same formulation as $M_1$ adding staying probability stratified by urbanisation level. $M_1$ + EC is the same formulation as $M_1$ adding conditional entropy stratified by urbanisation level. $M_1$ + IF is the same formulation as $M_1$ adding inflow stratified by urbanisation level. Maps only show the 670 districts modelled. This figure shows that mobility covariates have minimal impact on predictive skill at the subregion or district levels.

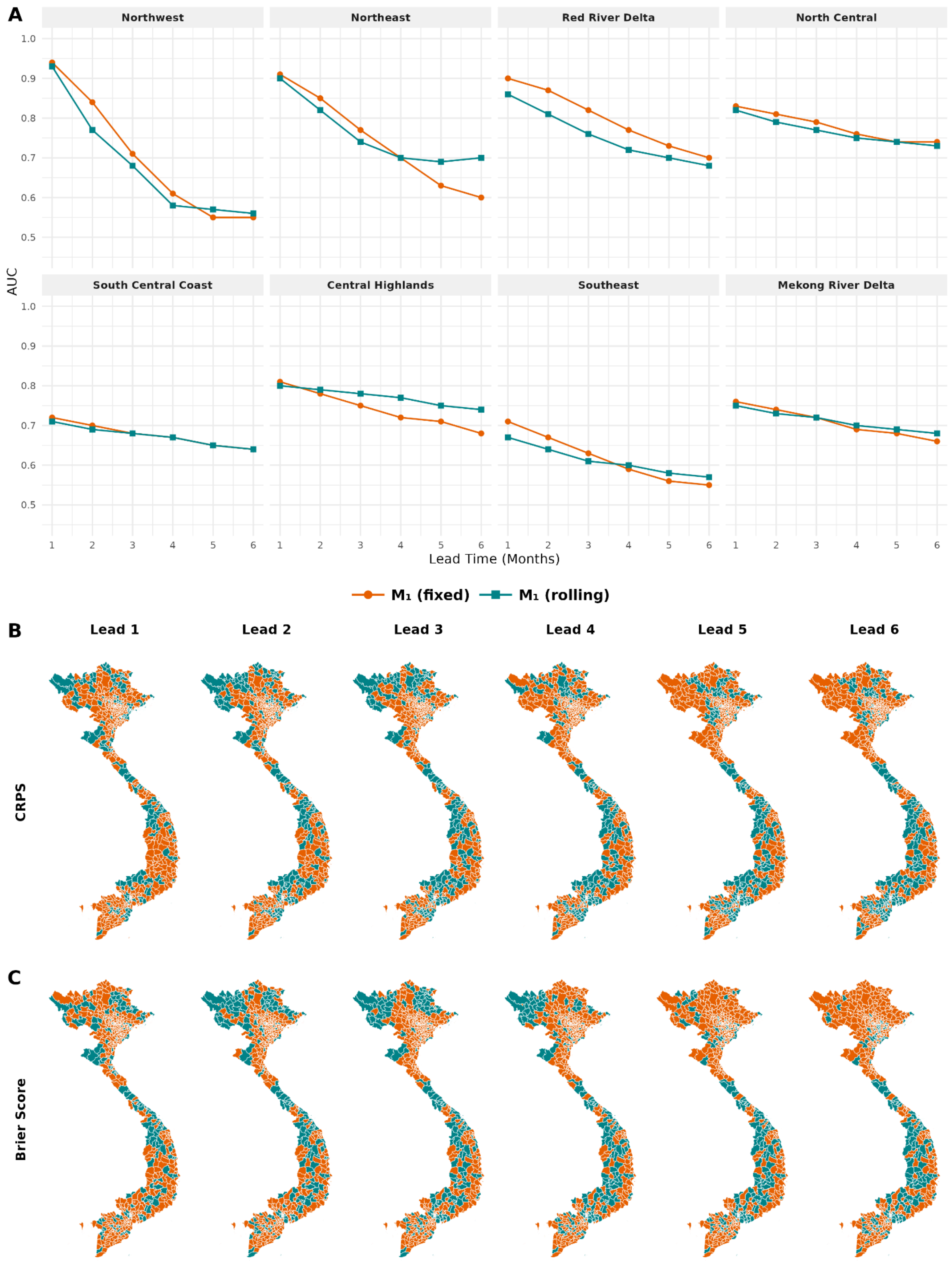


**Figure S12. Subregional and district-level predictive skill for derivations of model $M_1$ with a fixed vs rolling definition of the dengue season across lead times of 1 to 6 months.** (A) AUC by subregion and lead time for the two models. (B-C) Best-performing model per district by (B) CRPS and (C) Brier Score, at each lead time (columns) coloured by model. Fixed defines the dengue season consistently from May to April. Rolling dynamically defines the dengue season anchored such that the prediction month is the final month. Maps only show the 670 districts modelled.

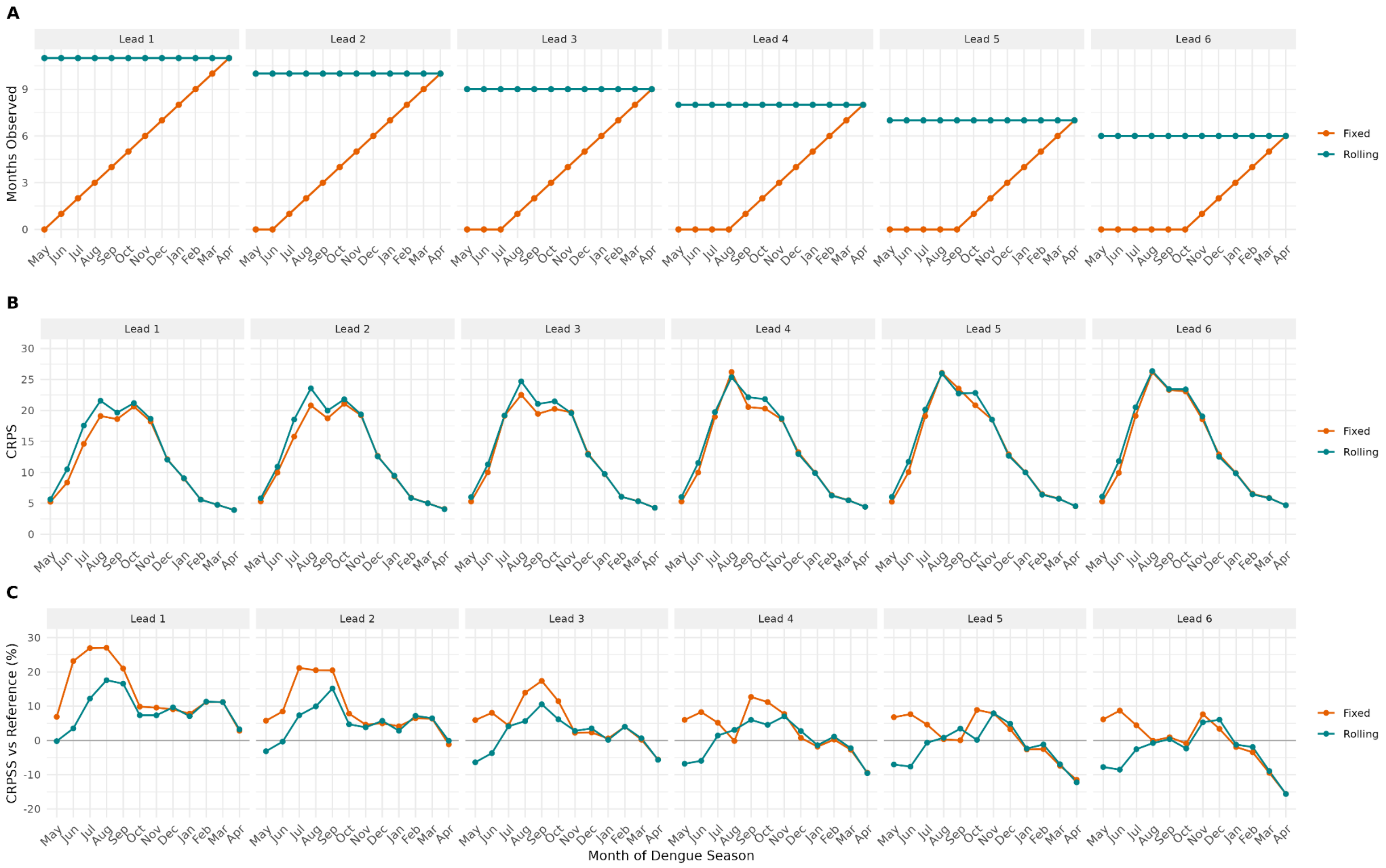


**Figure S13. Predictive skill by month of the dengue season for model $M_1$ with a fixed vs rolling year definition, Continuous Ranked Probability Score (CRPS).** (A) Number of months observed in the dengue season of the prediction month to inform the yearly random effect. (B) Mean CRPS by target month. (C) CRPS skill (CRPSS) relative to a seasonal reference, which uses case numbers from prior years of the same district-month as simulated predictions, where positive values indicate improvement over the reference. Fixed defines the dengue season consistently from May to April. Rolling dynamically defines the dengue season anchored such that the prediction month is the final month.

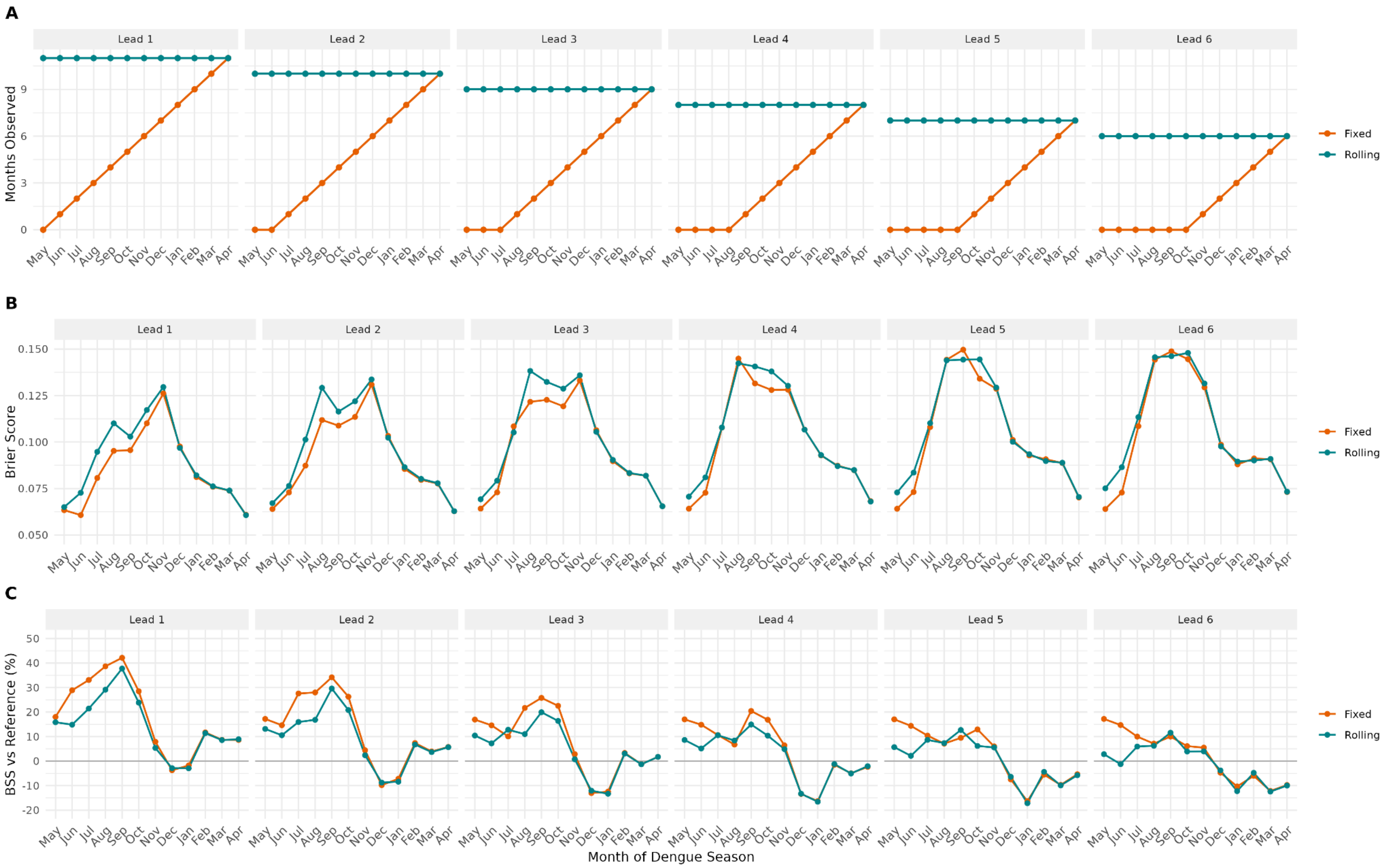


**Figure S14. Predictive skill by month of the dengue season for model $M_1$ with a fixed vs rolling year definition, Brier Score (BS).** (A) Number of months observed in the dengue season of the prediction month to inform the yearly random effect. (B) Mean BS by target month. (C) BS skill (BSS) relative to a seasonal reference, which issues as its forecast probability the proportion of previous years in which that district-month exceeded the outbreak threshold, where positive values indicate improvement over the reference. Fixed defines the dengue season consistently from May to April. Rolling dynamically defines the dengue season anchored such that the prediction month is the final month.

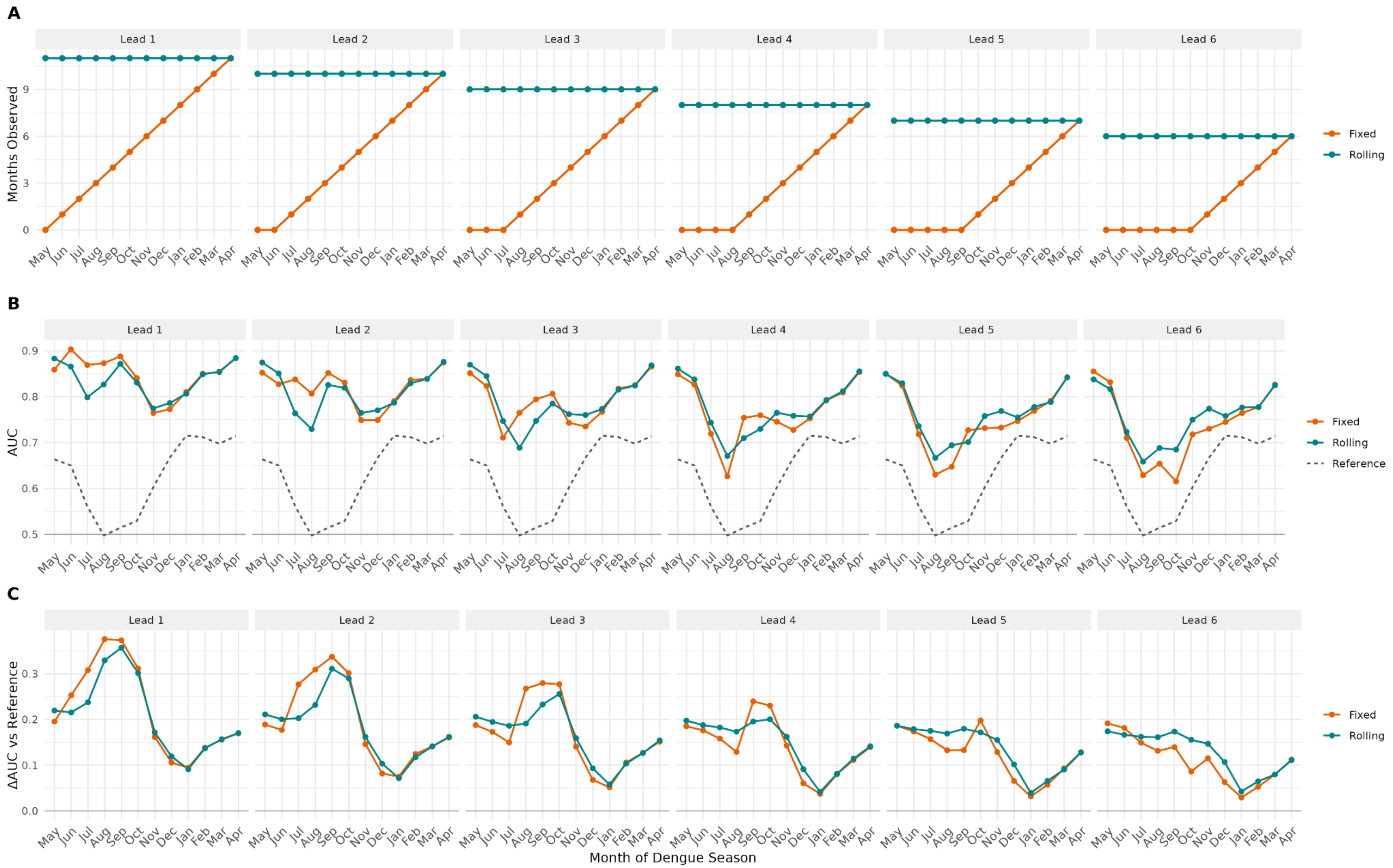


**Figure S15. Predictive skill by month of the dengue season for model $M_1$ with a fixed vs rolling year definition, Area Under the ROC Curve (AUC).** (A) Number of months observed in the dengue season of the prediction month to inform the yearly random effect. (B) AUC by target month. (C) Change in AUC (ΔAUC) relative to a seasonal reference, Reference, which ranks districts by the proportion of previous years in which that district-month exceeded the outbreak threshold, where positive values indicate better discrimination over the reference. Fixed defines the dengue season consistently from May to April. Rolling dynamically defines the dengue season anchored such that the prediction month is the final month.

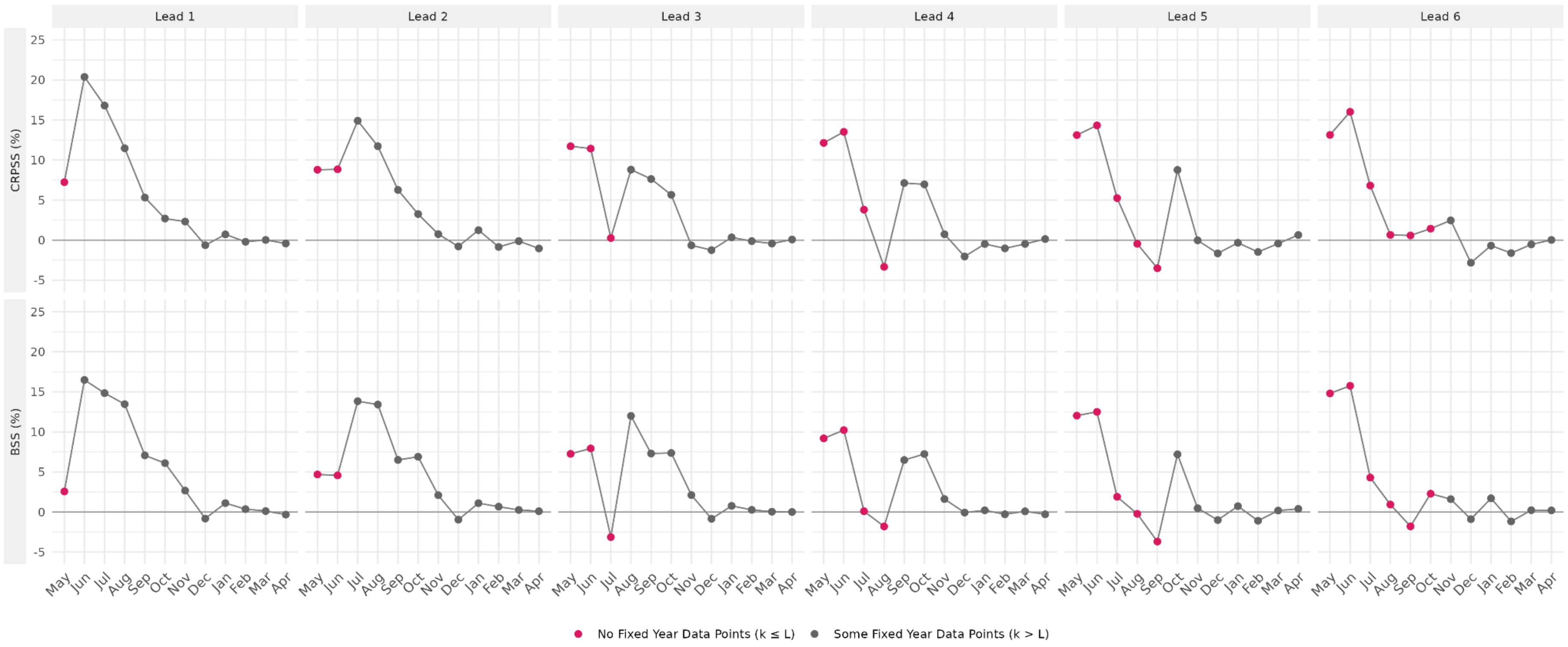


**Figure S16. Predictive skill by target month of model $M_1$ with a fixed year definition compared to a rolling year definition.** Continuous Ranked Probability Skill Score (CRPSS) and Brier Skill Score (BSS) computed from district-level scores per prediction month for model $M_1$ with a fixed year definition compared to a rolling year definition. Positive values indicate the fixed year performs better than the rolling year, and negative values show the inverse. Points are coloured by whether the fixed model had any observations from the dengue season of the prediction month to inform the yearly random effect, where k represents the position of the prediction month within the dengue season (k = 1 in May to k = 12 in April) and L represents the lead time, such that $k \leq L$ indicates no observations.

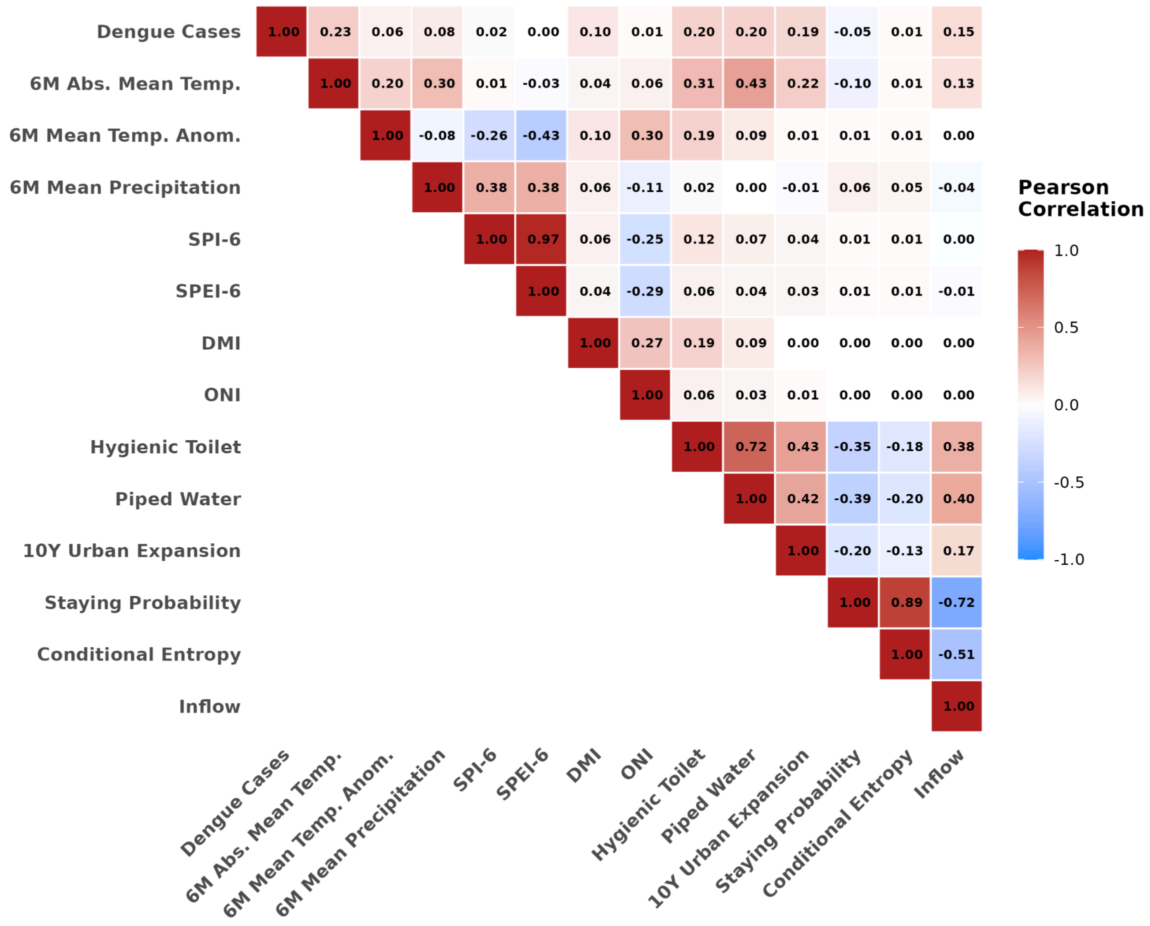


**Figure S17. Pairwise correlations between dengue cases and candidate covariates.** Pearson correlation coefficients between monthly district-level dengue case counts and a selection of climatic, oceanic, socioeconomic and mobility covariates across 670 districts in Vietnam, May 2001 to April 2021. Climatic variables are the 6-month absolute mean temperature, 6-month mean temperature anomaly, 6-month mean precipitation, 6-month Standardised Precipitation Index (SPI-6), and 6-month Standardised Precipitation Evapotranspiration Index (SPEI-6). Oceanic variables are the Dipole Mode Index (DMI) and Oceanic Niño Index (ONI). Climatic and oceanic variables are not lagged (0 months). Socioeconomic variables are the annual proportion of households per district with hygienic toilet access, the proportion with piped water access, and the 10-year urban expansion rate. Mobility variables are the staying probability, conditional outgoing entropy, and inflow, which are assumed static over time. Colour and value denote the direction and magnitude of the correlation. Only the upper triangle is shown to reduce redundancy.

**Table S1. Minimum and maximum contribution of compound climate extremes to dengue relative risk by subregion.** Values give the mean and 95% credible intervals for the dengue relative risk, $RR_{int}$, due to compound climate extremes in the best-fitting model. The interval is considered credible if it excludes 1. Compound extremes comprised a two-way interaction between the SPEI-6 lagged 5 months (Long SPEI-6), and SPEI-6 lagged 0 months (Short SPEI-6) stratified by subregion. The remaining covariate and random effects do not contribute to the relative risk. For each subregion, the minimum and maximum relative risks were evaluated using the 2.5th and 97.5th percentiles of each covariate observed per subregion. The covariate values corresponding to the minimum and maximum risks are stated.

| Subregion | Dengue Risk | $RR_{int}$ [95% CrI] | Long SPEI-6 | Short SPEI-6 |
|---|---|---|---|---|
| Northwest | Minimum | 0.69 [0.47, 1.02] | -1.86 | -1.86 |
| Northwest | Maximum | 1.75 [1.19, 2.59] | -1.86 | 1.39 |
| Northeast | Minimum | 0.33 [0.26, 0.41] | -1.91 | -1.91 |
| Northeast | Maximum | 1.56 [1.26, 1.89] | -1.91 | 1.40 |
| Red River Delta | Minimum | 0.61 [0.54, 0.71] | 1.37 | -1.89 |
| Red River Delta | Maximum | 2.53 [2.22, 2.83] | -1.89 | 1.42 |
| North Central | Minimum | 0.48 [0.42, 0.55] | 1.88 | 1.96 |
| North Central | Maximum | 1.34 [1.18, 1.50] | 1.88 | -1.72 |
| South Central Coast | Minimum | 0.55 [0.49, 0.61] | 1.82 | 1.85 |
| South Central Coast | Maximum | 2.08 [1.87, 2.31] | -1.81 | 1.85 |
| Central Highlands | Minimum | 0.65 [0.58, 0.75] | 1.87 | -1.87 |
| Central Highlands | Maximum | 1.45 [1.28, 1.65] | -1.87 | 1.89 |
| Southeast | Minimum | 0.67 [0.61, 0.73] | -1.91 | -1.91 |
| Southeast | Maximum | 1.40 [1.27, 1.56] | -1.91 | 1.79 |
| Mekong River Delta | Minimum | 0.62 [0.56, 0.66] | 1.82 | -1.88 |
| Mekong River Delta | Maximum | 1.74 [1.61, 1.88] | -1.88 | 1.81 |

**Table S2. Names and formulations of five models evaluated under rolling-origin cross-validation.** The predictive skill of five models at lead times of 1-6 months was evaluated under rolling-origin cross-validation each month from May 2016 to April 2021, including the best-fitting model ($M_1$), two models with the same covariate set ($M_2$, $M_3$), and two baseline random-effects models ($B_1$, $B_2$). The models estimate the log dengue incidence rate, $\mathbf{log(\rho_{s,t})}$, for district, **s**, in month, **t**. All formulations include a global intercept ($\boldsymbol{\alpha}$), monthly second-order random walk ($\boldsymbol{\delta_{m(t)}}$), and structured and unstructured spatial random effects specified as a modified Besag-York Mollié using a district-level adjacency matrix ($\mathbf{u_s + v_s}$). Formulations differ in whether they include a yearly independent and identically distributed random effect replicated by subregion ($\boldsymbol{\gamma_{r,a(t)}}$), whether they include climatic and socioeconomic covariates of the best fitting model ($\boldsymbol{\sum\beta_c X_c}$), and whether those covariates are estimated as random slopes stratified by subregion ($\mathbf{[...]_r}$). The specific covariates are: L = SPEI-6 (lagged 5 months), S = SPEI-6 (lagged 0 months), T = 6-month absolute mean temperature (lagged 0 months), D = Dipole Mode Index (lagged 4 months), HT = hygienic toilet access, UE = 10-year urban expansion rate, PW = piped water access.

| **Model** | **Monthly** ($\boldsymbol{\delta_{m(t)}}$) | **Yearly** ($\boldsymbol{\gamma_{r,a(t)}}$) | **Spatial** ($\mathbf{u_s + v_s}$) | **Covariates** ($\boldsymbol{\sum\beta_c X_c}$) | **Random Slopes** ($\mathbf{[...]_r}$) | **Equation** ($\mathbf{log(\rho_{s,t})}$) |
|---|---|---|---|---|---|---|
| $M_1$ | ✔ | ✔ | ✔ | ✔ | ✔ | $\alpha + \delta_{m(t)} + \gamma_{r,a(t)} + u_s + v_s + [\ \beta_L X_L + \beta_S X_S + \beta_{L,S} X_L X_S + \beta_T X_T + \beta_D X_D + \beta_{HT} X_{HT} + \beta_{UE} X_{UE} + \beta_{PW} X_{PW}\ ]_r$ |
| $M_2$ | ✔ | | ✔ | ✔ | ✔ | $\alpha + \delta_{m(t)} + u_s + v_s + [\ \beta_L X_L + \beta_S X_S + \beta_{L,S} X_L X_S + \beta_T X_T + \beta_D X_D + \beta_{HT} X_{HT} + \beta_{UE} X_{UE} + \beta_{PW} X_{PW}\ ]_r$ |
| $M_3$ | ✔ | ✔ | ✔ | ✔ | | $\alpha + \delta_{m(t)} + \gamma_{r,a(t)} + u_s + v_s + \beta_L X_L + \beta_S X_S + \beta_{L,S} X_L X_S + \beta_T X_T + \beta_D X_D + \beta_{HT} X_{HT} + \beta_{UE} X_{UE} + \beta_{PW} X_{PW}$ |
| $B_1$ | ✔ | ✔ | ✔ | | | $\alpha + \delta_{m(t)} + \gamma_{r,a(t)} + u_s + v_s$ |
| $B_2$ | ✔ | | ✔ | | | $\alpha + \delta_{m(t)} + u_s + v_s$ |

**Table S3. National predictive skill at 1-6-month leads.** National predictive metrics under rolling-origin cross-validation from May 2016 to April 2021. Models are $M_1$, $M_2$, $M_3$, $B_1$, and $B_2$ defined in Table S2. Metrics include the Continuous Ranked Probability Score (CRPS), Brier Score (BS), Area Under the ROC Curve (AUC), True Positive Rate (TPR), and False Positive Rate (FPR), with TPR and FPR calculated at optimal Trigger. Lower CRPS, BS and FPR, and higher AUC and TPR, indicate better performance.

| Model | Lead | CRPS | AUC | Trigger (%) | TPR (%) | FPR (%) | Brier Score |
|---|---|---|---|---|---|---|---|
| $M_1$ | 1 | 11.7 | 0.85 | 15 | 79 | 24 | 0.086 |
| $M_1$ | 2 | 12.3 | 0.82 | 12 | 78 | 26 | 0.092 |
| $M_1$ | 3 | 12.9 | 0.79 | 11 | 74 | 28 | 0.098 |
| $M_1$ | 4 | 13.3 | 0.76 | 10 | 70 | 30 | 0.102 |
| $M_1$ | 5 | 13.6 | 0.74 | 7 | 72 | 35 | 0.104 |
| $M_1$ | 6 | 13.8 | 0.72 | 7 | 70 | 36 | 0.105 |
| $M_2$ | 1 | 12.5 | 0.81 | 11 | 77 | 28 | 0.090 |
| $M_2$ | 2 | 12.7 | 0.79 | 10 | 74 | 29 | 0.093 |
| $M_2$ | 3 | 12.9 | 0.77 | 10 | 73 | 31 | 0.095 |
| $M_2$ | 4 | 13.0 | 0.76 | 9 | 71 | 31 | 0.097 |
| $M_2$ | 5 | 13.1 | 0.76 | 10 | 69 | 29 | 0.097 |
| $M_2$ | 6 | 13.1 | 0.75 | 9 | 70 | 31 | 0.098 |
| $M_3$ | 1 | 12.6 | 0.84 | 15 | 79 | 25 | 0.088 |
| $M_3$ | 2 | 13.1 | 0.82 | 12 | 76 | 28 | 0.094 |
| $M_3$ | 3 | 13.5 | 0.78 | 10 | 74 | 31 | 0.100 |
| $M_3$ | 4 | 13.7 | 0.75 | 10 | 68 | 31 | 0.104 |
| $M_3$ | 5 | 13.9 | 0.72 | 7 | 68 | 36 | 0.107 |
| $M_3$ | 6 | 13.9 | 0.71 | 5 | 70 | 40 | 0.109 |
| $B_1$ | 1 | 12.7 | 0.83 | 13 | 76 | 26 | 0.091 |
| $B_1$ | 2 | 13.3 | 0.79 | 10 | 73 | 30 | 0.098 |
| $B_1$ | 3 | 13.7 | 0.75 | 8 | 69 | 33 | 0.104 |
| $B_1$ | 4 | 13.9 | 0.71 | 6 | 67 | 37 | 0.109 |
| $B_1$ | 5 | 14.1 | 0.69 | 5 | 65 | 39 | 0.111 |
| $B_1$ | 6 | 14.2 | 0.67 | 3 | 67 | 43 | 0.112 |
| … | … | … | … | … | … | … | … |

| **Model** | **Lead** | **CRPS** | **AUC** | **Trigger (%)** | **TPR (%)** | **FPR (%)** | **Brier Score** |
|---|---|---|---|---|---|---|---|
| $B_2$ | 1 | 13.6 | 0.73 | 4 | 72 | 38 | 0.101 |
| $B_2$ | 2 | 13.6 | 0.72 | 4 | 70 | 38 | 0.101 |
| $B_2$ | 3 | 13.7 | 0.71 | 4 | 69 | 38 | 0.102 |
| $B_2$ | 4 | 13.7 | 0.70 | 3 | 71 | 41 | 0.102 |
| $B_2$ | 5 | 13.7 | 0.70 | 3 | 69 | 40 | 0.103 |
| $B_2$ | 6 | 13.7 | 0.70 | 3 | 71 | 42 | 0.103 |

**Table S4. Subregional predictive skill of $M_1$ relative to $B_1$ by lead time.** The Continuous Ranked Probability Skill Score (CRPSS) and Brier Skill Score (BSS) represent the percentage reduction in the mean Continuous Ranked Probability Score and Brier Score respectively, aggregated across districts within each subregion, per lead time. The Area Under the ROC Curve (AUC) difference gives the change in AUC per subregion per lead time. Positive values indicate improved skill for all metrics, and have been bolded. Columns represent lead times of 1-6 months per metric.

| | **CRPSS (%)** | | | | | | **BSS (%)** | | | | | | **AUC Difference (unitless)** | | | | | |
|---|---|---|---|---|---|---|---|---|---|---|---|---|---|---|---|---|---|---|
| **Subregion** | **Lead 1** | **Lead 2** | **Lead 3** | **Lead 4** | **Lead 5** | **Lead 6** | **Lead 1** | **Lead 2** | **Lead 3** | **Lead 4** | **Lead 5** | **Lead 6** | **Lead 1** | **Lead 2** | **Lead 3** | **Lead 4** | **Lead 5** | **Lead 6** |
| Northwest | -0.4 | -1.6 | **2.4** | **7.1** | **6.6** | **5.7** | **4.6** | **1.3** | **2.3** | **5.2** | **4.5** | **4.3** | 0.00 | **0.06** | **0.04** | 0.00 | -0.03 | -0.05 |
| Northeast | -17.3 | -21.0 | -19.7 | -13.8 | -9.5 | -8.2 | -8.7 | -11.2 | -12.3 | -11.5 | -8.9 | -7.3 | -0.02 | -0.02 | -0.04 | -0.08 | -0.12 | -0.14 |
| Red River Delta | **20.8** | **19.8** | **18.1** | **18.2** | **17.4** | **17.0** | **16.7** | **18.2** | **18.6** | **17.3** | **14.4** | **12.5** | **0.05** | **0.07** | **0.09** | **0.10** | **0.11** | **0.11** |
| North Central | -15.0 | -21.5 | -22.8 | -17.9 | -7.6 | 3.5 | -0.2 | -1.2 | -1.8 | -1.5 | **1.5** | **5.2** | **0.04** | **0.06** | **0.08** | **0.10** | **0.09** | **0.11** |
| South Central Coast | **10.7** | **10.0** | **8.9** | **7.0** | **6.6** | **8.1** | **7.2** | **8.0** | **8.6** | **9.0** | **9.1** | **10.3** | **0.04** | **0.05** | **0.05** | **0.06** | **0.06** | **0.06** |
| Central Highlands | **20.4** | **21.7** | **19.0** | **17.3** | **14.5** | **11.0** | **15.5** | **16.3** | **15.7** | **15.9** | **15.8** | **13.9** | **0.04** | **0.05** | **0.06** | **0.08** | **0.11** | **0.10** |
| Southeast | **3.2** | **1.0** | -2.0 | -4.8 | -8.6 | -11.6 | -1.7 | -3.3 | -4.4 | -4.6 | -6.1 | -7.3 | 0.00 | -0.01 | 0.00 | -0.01 | -0.02 | -0.01 |
| Mekong River Delta | **2.6** | **3.4** | **4.9** | **6.4** | **7.6** | **7.3** | **0.6** | **1.0** | **2.1** | **3.0** | **4.9** | **5.0** | **0.05** | **0.08** | **0.09** | **0.08** | **0.09** | **0.08** |

**Table S5. Subregional predictive skill of $M_2$ relative to $B_2$ by lead time.** The Continuous Ranked Probability Skill Score (CRPSS) and Brier Skill Score (BSS) represent the percentage reduction in the mean Continuous Ranked Probability Score and Brier Score respectively, aggregated across districts within each subregion, per lead time. The Area Under the ROC Curve (AUC) difference gives the change in AUC per subregion per lead time. Positive values indicate improved skill for all metrics, and have been bolded. Columns represent lead times of 1-6 months per metric.

| | **CRPSS (%)** | | | | | | **BSS (%)** | | | | | | **AUC Difference (unitless)** | | | | | |
|---|---|---|---|---|---|---|---|---|---|---|---|---|---|---|---|---|---|---|
| **Subregion** | **Lead 1** | **Lead 2** | **Lead 3** | **Lead 4** | **Lead 5** | **Lead 6** | **Lead 1** | **Lead 2** | **Lead 3** | **Lead 4** | **Lead 5** | **Lead 6** | **Lead 1** | **Lead 2** | **Lead 3** | **Lead 4** | **Lead 5** | **Lead 6** |
| Northwest | **8.2** | -1.9 | -6.0 | -5.9 | -8.3 | -8.5 | **4.8** | -12.9 | -20.1 | -19.1 | -21.2 | -21.7 | **0.19** | **0.08** | -0.05 | -0.08 | -0.09 | -0.10 |
| Northeast | **4.3** | -0.5 | -3.3 | -4.5 | -4.7 | -5.0 | **2.1** | -6.5 | -12.5 | -14.6 | -14.5 | -14.9 | **0.05** | -0.02 | -0.08 | -0.11 | -0.12 | -0.11 |
| Red River Delta | **11.9** | **8.2** | **6.9** | **5.8** | **5.3** | **5.2** | **10.2** | **3.8** | -0.4 | -2.0 | -2.5 | -3.0 | **0.10** | **0.08** | **0.07** | **0.07** | **0.06** | **0.06** |
| North Central | **10.7** | **9.2** | **9.2** | **8.6** | **7.8** | **6.2** | **12.1** | **8.6** | **7.6** | **6.0** | **5.7** | **4.9** | **0.05** | **0.04** | **0.05** | **0.04** | **0.05** | **0.05** |
| South Central Coast | **3.4** | **2.8** | **2.3** | **2.0** | **2.1** | **2.1** | **12.1** | **11.3** | **11.0** | **10.9** | **10.7** | **10.7** | **0.04** | **0.03** | **0.04** | **0.03** | **0.04** | **0.04** |
| Central Highlands | **14.9** | **14.5** | **13.6** | **12.8** | **12.9** | **12.7** | **19.6** | **19.0** | **18.4** | **18.0** | **18.0** | **17.9** | **0.10** | **0.11** | **0.11** | **0.12** | **0.12** | **0.12** |
| Southeast | **4.9** | **3.5** | **2.3** | **1.0** | **1.0** | **0.9** | **3.6** | **2.2** | **1.3** | **0.3** | **0.4** | **0.1** | -0.01 | -0.03 | -0.04 | -0.04 | -0.04 | -0.03 |
| Mekong River Delta | **10.2** | **9.4** | **9.3** | **9.1** | **9.0** | **9.0** | **9.3** | **8.8** | **8.8** | **8.9** | **9.3** | **9.3** | **0.06** | **0.05** | **0.04** | **0.05** | **0.04** | **0.04** |

**Table S6. Subregional predictive skill of $M_1$ relative to $M_2$ by lead time.** The Continuous Ranked Probability Skill Score (CRPSS) and Brier Skill Score (BSS) represent the percentage reduction in the mean Continuous Ranked Probability Score and Brier Score respectively, aggregated across districts within each subregion, per lead time. The Area Under the ROC Curve (AUC) difference gives the change in AUC per subregion per lead time. Positive values indicate improved skill for all metrics, and have been bolded. Columns represent lead times of 1-6 months per metric.

| | **CRPSS (%)** | | | | | | **BSS (%)** | | | | | | **AUC Difference (unitless)** | | | | | |
|---|---|---|---|---|---|---|---|---|---|---|---|---|---|---|---|---|---|---|
| **Subregion** | **Lead 1** | **Lead 2** | **Lead 3** | **Lead 4** | **Lead 5** | **Lead 6** | **Lead 1** | **Lead 2** | **Lead 3** | **Lead 4** | **Lead 5** | **Lead 6** | **Lead 1** | **Lead 2** | **Lead 3** | **Lead 4** | **Lead 5** | **Lead 6** |
| Northwest | **0.7** | **0.5** | **4.1** | **7.6** | **9.4** | **10.0** | **19.6** | **15.5** | **13.9** | **13.9** | **16.7** | **18.6** | **0.11** | **0.19** | **0.19** | **0.15** | **0.10** | **0.10** |
| Northeast | -16.9 | -24.4 | -23.9 | -17.3 | -12.9 | -10.6 | **7.0** | -3.4 | -10.4 | -12.0 | -8.7 | -3.5 | **0.08** | **0.13** | **0.12** | **0.09** | **0.03** | 0.00 |
| Red River Delta | **0.7** | -4.2 | -5.8 | -3.9 | -4.7 | -5.8 | **13.6** | **9.7** | **3.1** | -3.1 | -7.4 | -7.5 | **0.07** | **0.08** | **0.07** | **0.03** | 0.00 | -0.03 |
| North Central | -17.6 | -25.0 | -26.3 | -21.5 | -13.8 | -4.3 | -12.4 | -13.5 | -16.1 | -16.7 | -14.2 | -9.1 | **0.02** | **0.03** | **0.03** | **0.02** | 0.00 | **0.01** |
| South Central Coast | **2.8** | **0.3** | -1.0 | -1.4 | -0.7 | **1.0** | **0.6** | -1.4 | -3.4 | -4.4 | -5.9 | -5.8 | **0.01** | 0.00 | -0.02 | -0.02 | -0.04 | -0.05 |
| Central Highlands | **16.4** | **14.6** | **7.8** | **2.5** | -1.3 | -4.7 | **7.9** | **3.8** | -1.5 | -6.2 | -10.4 | -14.3 | **0.04** | **0.02** | 0.00 | -0.02 | -0.03 | -0.05 |
| Southeast | **13.2** | **9.4** | **4.4** | -0.7 | -6.2 | -10.6 | **9.9** | **5.1** | -0.2 | -3.9 | -8.0 | -9.7 | **0.08** | **0.06** | **0.03** | 0.00 | -0.03 | -0.04 |
| Mekong River Delta | **6.2** | **4.4** | **2.9** | **1.5** | -0.5 | -1.4 | **1.7** | -0.6 | -2.0 | -3.5 | -4.7 | -5.5 | **0.04** | **0.03** | **0.02** | -0.01 | -0.01 | -0.03 |

**Text S1. Fixed dengue season vs rolling dengue season model configurations for cross-validation analysis.**

The yearly random effect was fixed to a dengue season running from May to April to capture peaks within a single transmission cycle. Under a rolling-origin cross-validation design, the training window contains *max(0, k - L)* months of the dengue season for that prediction month at position *k* within that season ($k = 1$ in May to $k = 12$ in April) at lead time, $L$. This means that the within-season training data informing the yearly random effect varies systematically with prediction month. For example, the prediction is estimated from up to eleven months of within-season data for predictions in April, and from none whenever $k \leq L$, which at a 6-month lead time includes May to October.

We tested an alternative scheme with a rolling season definition, where the dengue season was dynamically defined to anchor the prediction month as the final (12th) month (Fletcher et al., 2025). Each season includes the prediction month and the eleven months preceding it such that the yearly effect is consistently informed by $12 - L$ observed months at every point in the season. Historical seasons containing fewer than 12 months at the start of the training data were removed prior to running rolling-origin cross-validation

As sensitivity analysis, we compared the best-fitting model, $M_1$, with a fixed year definition and rolling year definition. Predictive skill was quantified by the month of the dengue season to assess confounding with seasonality. Since the Continuous Ranked Probability Score (CRPS) and Brier Score (BS) scale with cases and outbreaks, respectively, both metrics track the seasonal cycle irrespective of model performance and are not directly comparable across months. Forecasts were additionally scored against a seasonal reference. For CRPS, the reference was the distribution of historical cases in that district-month. For BS, the reference was the proportion of preceding years in which the outbreak threshold was exceeded. For the Area Under the ROC Curve (AUC), the same proportion was used as the score for districts in each month. The skill of $M_1$ (fixed) and $M_1$ (rolling) relative to the seasonal reference was calculated as a percentage reduction in the CRPS (CRPSS, %) and the BS (BSS, %), as well as a difference in the AUC (ΔAUC). When computing the BS, month-lead combinations where no observations exceeded the threshold were excluded.

**Text S2. Mobility matrices and covariate descriptions and equations.**

We incorporated multiple formulations of spatial connectivity to determine whether metrics beyond adjacency of districts improve model performance both as additive covariates and random effects accounting for spatial structure.

***Spatial Connectivity Matrices***

First, we iteratively included spatial connectivity matrices to account for spatial autocorrelation across districts. Spatial dependency was modeled by iteratively incorporating distinct symmetric spatial connectivity matrices, beyond the standard neighborhood matrix (***W***), as spatially structured random effects within a modified Besag-York-Mollié (BYM2) framework. These matrices include: (i) distance-decay gravity models, (ii) travel time estimates developed by Gibb et al. (2023), and (iii) human mobility probabilities derived from Movement Range Maps (Meta, 2024). These are estimated as probabilities of mobility from each district to every other district (rowSum = 1), and are symmetric, whereby t(M) is the transposition of the mobility probability matrix (M):

*(M+t(M)) / 2 = M_symmetric*

***Gravity Model***

Human mobility between regions can be most easily approximated by accounting for population size and distance between regions. Two typical models are gravity models and radiation models. A gravity model attributes the attraction between two regions (*i* and *j*) to the population sizes (*P*) of them and repulsion between them as the distance (*d*).

$$Grav_{ij} = \frac{P_i P_j}{d_{ij}^{\gamma}}$$

This framework assumes that human mobility is driven completely by population size and distances neglecting other factors such as the presence of transport hubs, roads, as well as other sociodemographic and cultural factors. We constructed two distinct variations of the gravity model matrix. In both variations the baseline attraction between district i and district j ($i \neq j$) was modeled using the distance between the centroids of each i and j ($d_{ij}$) with a fixed distance decay parameter ($\gamma = 2$) and the population sizes *P* from the year 2021. In the first variation, self-loops were omitted by setting the diagonal elements to zero, and the off diagonal elements were symmetrized as required by the spatial random effects framework in R-INLA (Rue et al., 2009). In the second variation, we integrated the localised staying probability by extracting the diagonal elements of the human mobility probability matrix, representing the probability of individuals remaining in their home district, from the Movement Range Maps (Meta, 2024).

***Travel Time Estimates***

We accounted for travel time between districts rather than mere adjacency utilizing the travel time estimates from Gibb et al. (2023). The matrix we included is the population-weighted travel times with a manually adjusted travel time to offshore island districts. The travel times are the population weighted mean travel time from the centroid of district i, to all grid cells in district j. The diagonal for these matrices is either set to 0, or includes the mean travel time from the centroid of one district to all other grid cells in that same district. We also include a model in which we impose the time spent at home from the Movement Range matrix in the travel time diagonal.

***Movement Range Maps***

Human mobility measurements are direct measurements of the mobility of humans across space. This data is most typically available from call data records (CDR) or social media entities such as Google and Meta. While this data is imperfect, assuming that people logging into social media are representative of the whole population, previous work has shown that it is useful to inform our understanding of human movement (Belman et al., 2024;

Kiang et al., 2021; Salje et al., 2021). A key difference between gravity models and human mobility data is the approximation of time spent in one's home region. These can be approximated using the population sizes but human mobility data includes this element directly.

Here, we have downloaded movement distribution maps spanning 1 year (June 1, 2023 until June 1, 2024) from Meta AI For Good via the Humanitarian Data Exchange (Meta, 2024). These maps assign people to a geographic area (e.g. district) given their nighttime location. Each person is then assigned to a distance category using a random pinged location and its distance from home daily. These are then aggregated for each district to estimate the attributable fraction of movement from that district across 4 categories (0 km, 0-10 km, 10-100 km, 100+ km). We merged these attributable fractions with a stakeholder-provided district-level polygon shapefile for Vietnam, and reconciled any discordant districts in the GADM shapefile used by Facebook Data for Good. We transformed these location-specific-distance-fractions into an asymmetric matrix including the human mobility from every district to every other province or district by assigning each pair to one of the distance categories and dividing the distance fraction by the number of spatial units at each. We then symmetrized the matrix to align with the requirements for the BYM2 framework in R-INLA (Rue et al., 2009). We evaluated the performance of this human mobility matrix alone as well as imposing the diagonal (time spent in home region) from this matrix into the gravity models and travel time models (which do not explicitly account for time spent at home).

***Mobility Covariates***

We also developed three refined human mobility indicators from the Meta AI For Good Movement Range Maps to better capture transmission dynamics.

***Mobility Inflow***

The first covariate, normalized inflow ($\mathcal{I}_j$), quantifies the relative volume of individuals arriving into the district $j$ from all other origins. We first calculate the Expected Raw Inflow ($F_j$) as the sum of individuals moving from all origin districts $i$ to destination $j$, weighted by the origin population ($P_i$):

$$F_j = \sum_{i \neq j} M_{ij} P_i$$

We then normalized ($F_j$) by the maximum inflow observed across all districts to produce the final covariate:

$$\text{Inflow}_j = \frac{F_j}{\max_k(F_k)}$$

Where $M_{ij}$ represents the probability of movement from district $i$ to district $j$ in the asymmetric origin-destination matrix ($M$). By setting $i \neq j$, we ensure the metric reflects only mobility rather than local residence.

***Stay Probability***

The staying probability ($S_i$) represents the proportion of individuals who remain within their home district during the observed period. This is captured by the diagonal elements of the mobility matrix:

$$S_i = M_{ii}$$

*Conditional Outgoing Entropy*

To measure the diversity of travel destinations, we calculate the Conditional Shannon Mobility Entropy ($H_i^*$). This metric specifically describes the geographical spread of people leaving a district excluding those who are staying $M_{ii}$.

We first calculate the conditional transition probability ($M_{ij}^*$), which re-scales the probabilities so that movement to all other districts sums to 1:

$$M_{ij}^* = \frac{M_{ij}}{\sum_{k \neq i} M_{ik}}, \quad \text{for } j \neq i$$

We then calculate the entropy based on this conditional distribution:

$$H_i^* = -\sum_{j \neq i} M_{ij}^* \ln(M_{ij}^*)$$

A high $H_i^*$ indicates that travelers from district $i$ disperse to many different locations, whereas a low value suggests movement is concentrated toward a few specific destinations.

**References**


Belman, S., Lefrancq, N., Nzenze, S., Downs, S., du Plessis, M., Lo, S. W., McGee, L., Madhi, S. A., von Gottberg, A., Bentley, S. D., & Salje, H. (2024). Geographical migration and fitness dynamics of Streptococcus pneumoniae. *Nature*, *631*(8020), 386–392. https://doi.org/10.1038/s41586-024-07626-3

Fletcher, C., Moirano, G., Alcayna, T., Rollock, L., Van Meerbeeck, C. J., Mahon, R., Trotman, A., Boodram, L.-L., Browne, T., Best, S., Lührsen, D., Diaz, A. R., Dunbar, W., Lippi, C. A., Ryan, S. J., Colón-González, F. J., Stewart-Ibarra, A. M., & Lowe, R. (2025). Compound and cascading effects of climatic extremes on dengue outbreak risk in the Caribbean: An impact-based modelling framework with long-lag and short-lag interactions. *The Lancet Planetary Health*, *9*(8), 101279. https://doi.org/10.1016/j.lanplh.2025.06.003

Gibb, R., Colón-González, F. J., Lan, P. T., Huong, P. T., Nam, V. S., Duoc, V. T., Hung, D. T., Dong, N. T., Chien, V. C., Trang, L. T. T., Kien Quoc, D., Hoa, T. M., Tai, N. H., Hang, T. T., Tsarouchi, G., Ainscoe, E., Harpham, Q., Hofmann, B., Lumbroso, D., … Lowe, R. (2023). Interactions between climate change, urban infrastructure and mobility are driving dengue emergence in Vietnam. *Nature Communications*, *14*(1), 8179. https://doi.org/10.1038/s41467-023-43954-0

Kiang, M. V., Santillana, M., Chen, J. T., Onnela, J.-P., Krieger, N., Engø-Monsen, K., Ekapirat, N., Areechokchai, D., Prempree, P., Maude, R. J., & Buckee, C. O. (2021). Incorporating human mobility data improves forecasts of Dengue fever in Thailand. *Scientific Reports*, *11*(1), 923. https://doi.org/10.1038/s41598-020-79438-0

Meta. (2024). *Movement Distribution Maps | AFG Dataset | AI at Meta*. https://ai.meta.com/ai-for-good/datasets/movement-distribution-maps/

Rue, H., Martino, S., & Chopin, N. (2009). Approximate Bayesian Inference for Latent Gaussian models by using Integrated Nested Laplace Approximations. *Journal of the Royal Statistical Society Series B: Statistical Methodology*, *71*(2), 319–392. https://doi.org/10.1111/j.1467-9868.2008.00700.x

Salje, H., Wesolowski, A., Brown, T. S., Kiang, M. V., Berry, I. M., Lefrancq, N., Fernandez, S., Jarman, R. G., Ruchusatsawat, K., Iamsirithaworn, S., Vandepitte, W. P., Suntarattiwong, P., Read, J. M., Klungthong, C., Thaisomboonsuk, B., Engø-Monsen, K., Buckee, C., Cauchemez, S., & Cummings, D. A. T. (2021). Reconstructing unseen transmission events to infer dengue dynamics from viral sequences. *Nature Communications*, *12*(1), 1810. https://doi.org/10.1038/s41467-021-21888-9
